# DNA methylation-wide association study of prevalent and incident dementia in the US Health and Retirement Study

**DOI:** 10.1101/2025.11.10.25339919

**Authors:** John Dou, Scarlet Cockell, Herong Wang, Nathan Hemenway, Lindsay Ryan, Matt Zawistowski, Erin B. Ware, Kelly M. Bakulski

## Abstract

**BACKGROUND:** Peripheral blood DNA methylation may have utility as an early dementia risk biomarker.

**METHODS:** We analyzed DNA methylation (blood collected 2016) and cognitive impairment in the Health and Retirement Study, a longitudinal study representative of US adults over age 50 (3,921 individuals and 585,356 CpG sites). We analyzed methylation associations with cognitive status both cross-sectionally and prospectively among participants with normal cognition at baseline with four years follow-up.

**RESULTS:** Cross-sectionally, 5,322 CpGs were associated (p-value<0.01) with cognitive impairment non-dementia, and 14,366 (166 genome-wide FDR<0.05) with dementia. Prospectively, 4,898 CpGs were associated with any-impairment. Enriched biologic pathways include ion transport, ligand-gated channel, and neuron differentiation. Nine CpGs overlapped all analyses including cg02583484 (*HNRNPA1*), cg15266133 (*LOC102724084*), cg24287460 (*CCDC48*), cg17124509 (*C17orf57*), and cg02553054 (*SMARCD1*).

**DISCUSSION:** CpGs identified were enriched in pathways related to Alzheimer’s disease pathology and provide promising grounds for non-invasive blood biomarkers. Future studies for replication and with longer follow-up are needed.

## 1 BACKGROUND

Dementia is a progressive neurodegenerative disease which impacts a person’s ability to think, remember, and carry out activities of daily life.^1^ Roughly 10 million new dementia cases are reported yearly, with incidence anticipated to reach 13.8 million by 2060 without the development of novel prevention or treatment measures.^2–5^ Dementia is often experienced along a clinical continuum starting at the asymptomatic preclinical phase, passing through the prodromal stage of mild cognitive impairment, and ending in dementia.^6,7^ The path to clinical diagnosis of dementia is indirect and involves a combination of neurological exams, brain imaging, and cerebrospinal fluid or blood tests to ensure accuracy of diagnosis.^2,6^ Outstanding recent advances in blood biomarkers have improved Alzheimer’s disease diagnostics in research settings, and clinical applications are ongoing.^8^ Building on these successes, the field of dementia research faces an urgent need for susceptibility and risk biomarkers reflecting earlier stages in the disease processes as well as easily available and inexpensive diagnostic tools for dementia more broadly to increase the possibility of early detection, diagnosis, and intervention.^2,6^ Current research focuses on using epigenetic markers, such as DNA methylation, to identify signatures in peripheral blood associated with dementia which may have future utility as biomarkers.^6,9,10^

Epigenetics, the study of reversable modifications to DNA which alter gene expression in response to environmental or behavioral factors, contribute to the development of numerous human pathologies.^6^ The most commonly studied epigenetic marker is DNA methylation, which refers to a cytosine nucleotide covalently bonded to a methyl group upstream of a guanine (CpG).^11,12^ DNA methylation markers can act mechanistically to influence gene regulation and can have research utility as biomarkers of prior exposure or precursors of future disease.^11^ In fact, studies have identified blood based DNA methylation differences in genes associated with the diagnosis of Alzheimer’s disease such as, *SPIDR and CDH6*.^13,14^ Other studies investigating candidate biomarkers in the blood of participants with early dementia have found mixed results.^6,9,13,15,16^ The long duration of time between pathological changes and symptom onset provides challenges in comprehensive characterization of biomarkers.^13^ However, longitudinal studies leveraging diverse demographic and genetic cohorts, well powered to interrogate associations, aid in further characterization of blood based biomarkers for dementia.

In this analysis, we conducted epigenome-wide association studies of blood DNA methylation with cognitive status in the United States Health and Retirement Study (HRS) cohort (waves 2016-2020). We separately tested DNA methylation for associations with prevalent and incident cognitive status. Among 3,395 participants from the 2016 wave of the HRS we evaluated cross sectional associations of site-specific DNA methylation and prevalent cognitive function. Among 2,424 participants in the 2016-2020 waves of the HRS, we assessed associations of baseline DNA methylation and incident cognitive status. We conducted gene set enrichment analyses to identify biological pathways significantly enriched among prioritized DNA methylation sites. Considering sample size, longitudinal outcome data, and a diverse cohort, this study advances knowledge of blood-based DNA methylation biomarkers of dementia.

## 2 METHODS

### 2.1 Study population

The HRS is a longitudinal panel cohort study that began in 1992, and includes a nationally representative sample of participants 50 years and older.^17^ Every two years, participants are assessed for sociodemographic, health (including cognitive status), and financial (among other areas) experiences.^17^ A detailed description of this study has been given previously.^17^ Participants provided written informed consent at the time of participation. This secondary data analysis was approved by the University of Michigan Institutional Review Board (HUM00128220). Our prospective analyses used DNA methylation data collected from venous blood in 2016 and cognition data collected from survey measures in 2016. The incident analyses used DNA methylation from 2016, and cognition data from 2018 and 2020 (**Figure 1**). Demographic and cognitive data are publicly available (https://hrs.isr.umich.edu).

**Figure 1.**
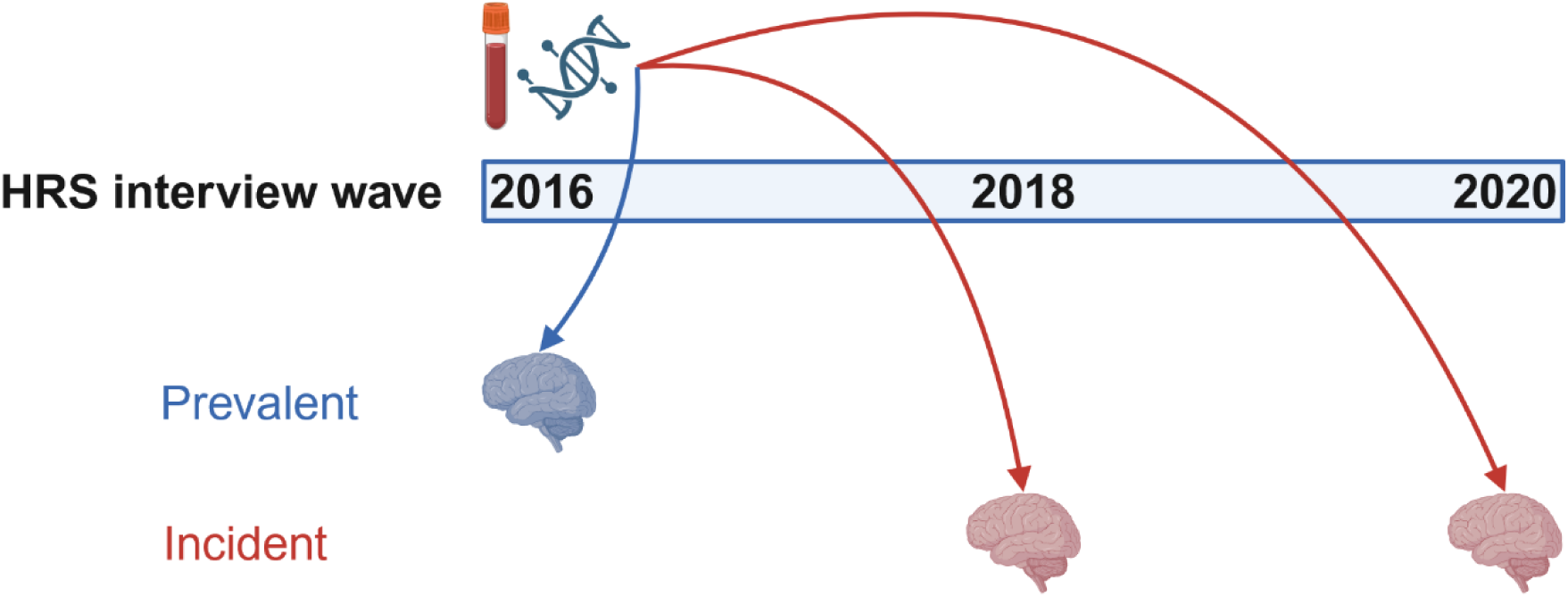
Study design timing. DNA methylation (DNAm) was collected once in 2016. Cognitive assessments were performed in 2016, 2018, and 2020. Associations of single site DNAm and prevalent cognitive function were assessed in the 2016 interview wave (N = 3,395). Associations between DNAm and incident cognitive status were assessed in the 2018-2020 interview waves (N = 2,424).

### 2.2 DNA methylation measures

In 2016, the HRS Venous Blood Study added numerous biomarker measures, including DNA methylation using the Infinium Methylation EPIC BeadChip v1.0.^18^ DNA methylation data were accessed through the National Institute on Aging Genetics of Alzheimer’s Disease Storage (NIAGADS) site (dataset ng000153; https://dss.niagads.org/datasets/ng00153/).

### 2.3 DNA methylation data pre-processing

The HRS conducted initial sample filtering of DNA methylation data. In brief, DNA methylation data was obtained on 4,224 samples, including 81 cell line controls and 40 blinded duplicates. The following samples were removed: controls, a blinded duplicate pair that did not have correlated methylation values, samples with >5% of probes failing detection-p (*minfi* method, p>0.01), and sex mismatched samples. After these sample filtering criteria, red-green channel set raw data were released on 4,018 samples. Full details can be found on documentation on the HRS website: https://hrs.isr.umich.edu/sites/default/files/genetic/HRS_DNAm_OCT2023.pdf.

Our study team performed additional quality control checks on the released 4,018 methylation samples using the *ewastools* package.^19^ We used a more stringent p-value threshold of p>0.0001 for detection-p checks. A total of 95,491 probes that failed detection-p in >1% of samples were removed. An additional 38,904 probes were removed for being previously flagged as having mapping or cross-reactivity problems (**Supplemental Figure 1**).^10^

We also applied additional quality control criteria for filtering samples. We removed 10 samples that both failed an Illumina control probe metric and had low intensity values (defined as sum of median log2 intensity methylated and unmethylated signal < 20). We confirmed that there were no sex mismatched samples remaining.

The DNA methylation array contains 59 single nucleotide polymorphism (SNP) probes, which we used to evaluate sample identity. Because these DNA methylation samples were expected to be from unique, unrelated, participants, we first calculated between-sample SNP probe Pearson correlations. Second, we evaluated sample identity by comparing the DNA methylation array SNP probes to existing genotyping array data on these samples^17^ (data available from the National Institute on Aging Genetics of Alzheimer’s Disease Data Storage Site: NG00153) using Pearson correlations. We observed unexpectedly high levels of correlation (SNP probe correlations >0.75) for 18 samples (9 pairs), where identity based on genotyping samples could not be clearly determined and these samples were dropped. In these samples, there were either no genetic data to compare against, or correlation between methylation array and genotype array SNPs were ambiguous (correlation<0.9 for all DNAm probe SNP and genotype array SNP pairwise comparison, or correlation high for both pairwise comparisons). Additionally, in five pairs of samples the true identity of samples were able to be determined from matching with genotyping data, resulting in five samples dropped, with the other in the pair retained. In total, 23 samples were dropped after examining SNP-probe correlation.

We next removed 59 samples with >0.95% of probes failing detection-p. Additionally, we assessed cell composition and compared to expected distributions in adult blood samples. ^20^ We removed 5 samples with outlier estimated granulocyte (<5%) proportion. In total, we removed 97 samples (**Supplemental Figure 1**). Informed by existing evaluations of DNA methylation processing methods^21^, DNA methylation data normalization was done with a combination of dye-bias and noob background correction^22^ followed by BMIQ normalization.^23^ To focus our analyses on the sites with the greatest variability in our sample, we further removed CpG sites with the 20% lowest variance, roughly corresponding to variance < 0.001. Thus, our final DNA methylation data matrix was 3,921 samples and 585,356 CpG sites.

### 2.4 Cognitive measures

For cognitive status measures, we used the HRS Cross-Wave Imputation of Cognitive Functioning Measures for the years 2016 to 2020 with categorization into three levels based on the Langa-Weir classification (normal, CIND, dementia).^24^ Self-respondent cognitive status was assessed on a 27-point scale by summing an immediate and delayed 10-noun free recall test (0 to 20 points), a serial 7 subtraction test (0 to 5 points), and a backward count from 20 test (0 to 2 points).^17,25^ Respondents were classified into normal cognition (score range: 12-27), cognitive impairment non-dementia (CIND) (7-11), and dementia (0-6).^25^ For respondents represented by a proxy (baseline N= 153, follow-up N= 812), an alternative 11-point scale was used.^25^ The proxy’s assessment of the respondent’s memory ranged from excellent to poor (0 to 4 points), the number of instrumental activities of daily living with limitations (0 to 5 points), and the survey interviewer’s assessment of whether the respondent had difficulty completing the interview because of a cognitive limitation (0 to 2 points).^25^ Established cutoffs based on proxy scores classified the respondent into one of the three categories: normal cognition (0-2), CIND (3-5), or dementia (6-11).^25^

For prevalent analyses, we used the three category cognitive status outcome variable (normal, CIND, dementia)with normal cognition as the reference group. In the incident analyses, for sample size purposes, we created a two category cognitive status outcome variable (normal, any cognitive impairment) by combining the “dementia” and “CIND” categories into one category of “any cognitive impairment”, normal cognition was used as the reference group.

### 2.5 Covariate measures

Baseline (2016) participant sociodemographic measures were provided by self-reported questionnaire. These include age (years), participant sex (male, female), race/ethnicity (non-Hispanic white, non-Hispanic Black, Hispanic, non-Hispanic Other), educational attainment (years), smoking status (current, former, never), and alcohol consumption (#days/week).

Participant age was calculated based on the difference between birth date and visit date. Body mass index (kg/m^2^) was calculated based on height and weight. Apolipoprotein E ε4 allele (*APOE-ε4*) carrier status (yes/no for having any copy ε4 allele), measured by Taqman allelic discrimination SNP assay or imputed from genotype array data, was downloaded from The National Institute on Aging Genetics of Alzheimer’s Disease Data Storage Site (NIAGADS, Insert: accession number) (https://dss.niagads.org/sample-sets/snd10040/). We computed two sets of covariates derived from methylation: estimated cell types and ancestry principal components. Using the *MethylGenotyper* package^26^, which utilizes intensities of probes near SNPs, we computed four ancestry principal components from the methylation data. We generated estimates for cell type proportions (granulocytes, natural killer cell, B cell, CD4+ T cell, CD8+ T cell, monocytes) using the Salas reference panel^27^ with the Houseman deconvolution method.^28^

### 2.6 Statistical analyses

All analyses were conducted in R statistical software (version 4.3).^29^ An independent analyst conducted code review and code to produce all analyses is available from our github page (https://github.com/bakulskilab/HRS-DNAm-cognition).

Participants were eligible for the prevalent cognitive impairment analysis if there was complete information on DNA methylation, key covariates described above, and cognitive status at baseline. The incident sample was a subset who had normal cognition at baseline and who had at least one subsequent cognitive measure (in either 2018 or 2020). We visualized participant inclusion and exclusion using a flow chart. We compared the distributions of variables between the excluded and included samples using mean and standard deviation for continuous measures and count and frequency for categorical measures. Within the included samples, we compared the distributions of covariates by cognitive status.

In prevalent analyses, to perform an epigenome-wide association study of site-specific DNA methylation and cognition, we fit parallel linear regression models for each site using the *limma* package.^30^ We tested for association with prevalent cognitive status, modeled as a three level categorical variable of cognitively impaired non-dementia, dementia, and normal cognition as the reference group. Site-specific models were adjusted for age, sex, years of education, proportions of cell types (leaving out monocytes), and ancestry principal components. A health behavior sensitivity model additionally adjusted for body mass index, smoking status, and alcohol use. Finally, an *APOE* sensitivity model adjusted for *APOE-ε4* carrier status, in a smaller sample restricted to those with genotype data.

In incident analyses, models for cognitive impairment were fit in similar manner, restricted to those cognitively normal at baseline. We investigated the association between DNA methylation and incident any cognitive impairment and compared to those retaining normal cognition. Models were adjusted for the same covariates as the prevalent model, but with follow up time (number of years since 2016) as an additional variable. The same health behavior and *APOE* sensitivity models as for the prevalent models were fit for the any incident cognitive impairment model. Additionally, for the incident sample, we performed exploratory analysis using the three category cognition outcome (CIND, dementia, normal cognition) to ascertain associations between DNAm and incident CIND and incident dementia as separate categories.

For all single site models, we used the same visualization and diagnostic analysis approaches. We evaluated single site model performance by creating Q-Q plots and computing lambda values. We considered sites with p-values < 0.01 nominally associated. We accounted for multiple comparisons with the Benjamini-Hochberg method to calculate false discovery rate (FDR) adjusted p-values.^31^ In tables, we reported the magnitude of association as the adjusted percent difference in DNA methylation in the impaired group of interest versus normal cognition, the 95% confidence interval, the unadjusted p-value, and the FDR. We visualized results across all models using volcano plots of the magnitude of the association and the level of significance. We visualized the results at the top 10 sites using boxplots of methylation values by group. We compared results across models using a correlation matrix of effect estimates and examined overlap of nominally associated sites with UpSet plots.^32^

To assess whether the DNA methylation sites differentially methylated by cognitive status were enriched for biologic pathways, we performed gene set enrichment analysis using the *methylGSA* package.^33^ We used the GSEAPreranked approach using the methylRRA function. To understand the genomic context of our findings, we examined whether differentially methylated sites were enriched in CpG island regions (islands, shores, shelves, open sea). We also analyzed the differentially methylated sites for enrichment of tissue specific chromatin state marks using the eFORGE tool,^34^ using the top 1000 CpGs by p-value as input.

For replication testing, we compared our results to published epigenome-wide association studies, prioritizing the largest existing studies conducted in venous blood with available summary statistics. Roubroeks 2020 examined peripheral blood DNA methylation in 284 individuals (89 controls, 86 Alzheimer’s disease, 109 mild cognitive impairment), and had summary statistics released for top 1000 CpGs for Alzheimer’s disease vs control, and top 1000 CpGs for mild cognitive impairment vs control.^35^ Sommerer 2022 examined peripheral blood DNA methylation in relation to episodic memory in 1019 individuals cross-sectionally, and 626 individuals longitudinally, with full summary statistics for all CpGs tested released.^36^ We evaluated overlapping CpGs with nominal p-value < 0.01 in our study and the Sommerer 2022 study. Since Roubroeks 2020 only had summary statistics for the top 1000 CpGs, overlap was evaluated between those top 1000 and the top CpGs (nominal p-value < 0.01) in our study. Number of overlapping CpGs were visualized using UpSet plots.

## 3 RESULTS

### 3.1 Sample descriptives

After filtering for methylation quality control and missing covariates, 3,395 participants were included in the primary prevalent cognitive impairment analysis and 2,703 participants for the incident cognitive impairment analysis (**Supplemental Figure 2**). The *APOE* sensitivity model was restricted to 3,370 participants in prevalent analysis and 2,401 participants in incident analysis. Excluded participants were younger on average, more likely to be Hispanic or non-Hispanic Black or Other, and more likely to be current smokers (**Supplementary Table 1, Supplementary Table 2**).

Among the primary prevalent analytic sample there were 651 (17.0%) individuals categorized as CIND, and 145 (3.8%?) as dementia. Cognitively normal individuals were younger at baseline, more likely to be non-Hispanic White, have more years of education, more likely to be never smokers, and more likely to have no copies of the *APOE-ε4* allele (**Table 1**).

**Table 1.**
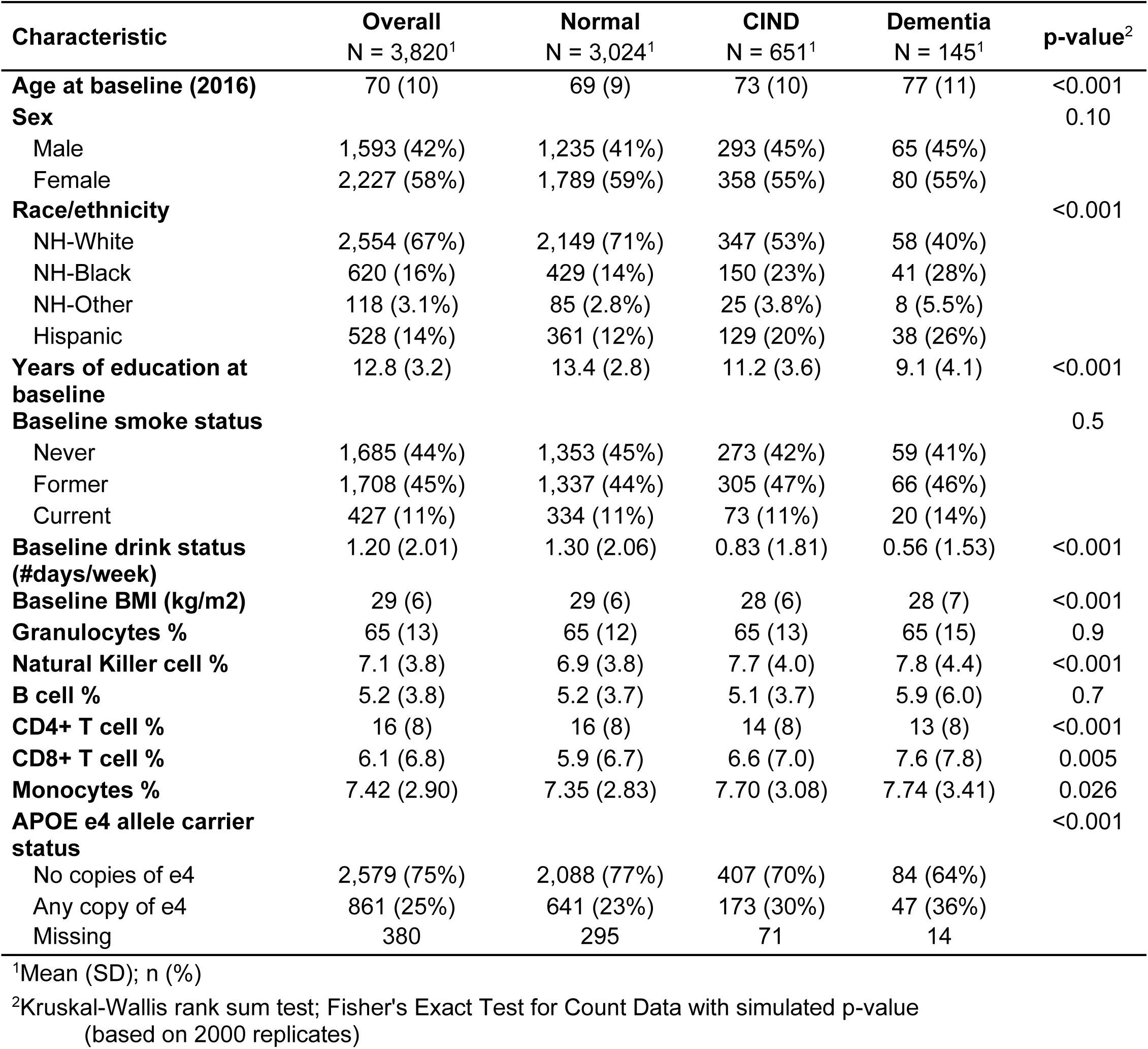
Descriptive statistics for participants included in the prevalent analyses, by prevalent cognitive status: normal, cognitively impaired non-dementia (CIND), dementia.

| **Characteristic** | **Overall**<br>**N = 3,820<sup>1</sup>** | **Normal**<br>**N = 3,024<sup>1</sup>** | **CIND**<br>**N = 651<sup>1</sup>** | **Dementia**<br>**N = 145<sup>1</sup>** | **p-value<sup>2</sup>** |
| --- | --- | --- | --- | --- | --- |
| **Age at baseline (2016)** | 70 (10) | 69 (9) | 73 (10) | 77 (11) | <0.001 |
| **Sex** |  |  |  |  | 0.10 |
| Male | 1,593 (42%) | 1,235 (41%) | 293 (45%) | 65 (45%) |  |
| Female | 2,227 (58%) | 1,789 (59%) | 358 (55%) | 80 (55%) |  |
| **Race/ethnicity** |  |  |  |  | <0.001 |
| NH-White | 2,554 (67%) | 2,149 (71%) | 347 (53%) | 58 (40%) |  |
| NH-Black | 620 (16%) | 429 (14%) | 150 (23%) | 41 (28%) |  |
| NH-Other | 118 (3.1%) | 85 (2.8%) | 25 (3.8%) | 8 (5.5%) |  |
| Hispanic | 528 (14%) | 361 (12%) | 129 (20%) | 38 (26%) |  |
| **Years of education at baseline** | 12.8 (3.2) | 13.4 (2.8) | 11.2 (3.6) | 9.1 (4.1) | <0.001 |
| **Baseline smoke status** |  |  |  |  | 0.5 |
| Never | 1,685 (44%) | 1,353 (45%) | 273 (42%) | 59 (41%) |  |
| Former | 1,708 (45%) | 1,337 (44%) | 305 (47%) | 66 (46%) |  |
| Current | 427 (11%) | 334 (11%) | 73 (11%) | 20 (14%) |  |
| **Baseline drink status (#days/week)** | 1.20 (2.01) | 1.30 (2.06) | 0.83 (1.81) | 0.56 (1.53) | <0.001 |
| **Baseline BMI (kg/m2)** | 29 (6) | 29 (6) | 28 (6) | 28 (7) | <0.001 |
| **Granulocytes %** | 65 (13) | 65 (12) | 65 (13) | 65 (15) | 0.9 |
| **Natural Killer cell %** | 7.1 (3.8) | 6.9 (3.8) | 7.7 (4.0) | 7.8 (4.4) | <0.001 |
| **B cell %** | 5.2 (3.8) | 5.2 (3.7) | 5.1 (3.7) | 5.9 (6.0) | 0.7 |
| **CD4+ T cell %** | 16 (8) | 16 (8) | 14 (8) | 13 (8) | <0.001 |
| **CD8+ T cell %** | 6.1 (6.8) | 5.9 (6.7) | 6.6 (7.0) | 7.6 (7.8) | 0.005 |
| **Monocytes %** | 7.42 (2.90) | 7.35 (2.83) | 7.70 (3.08) | 7.74 (3.41) | 0.026 |
| **APOE e4 allele carrier status** |  |  |  |  | <0.001 |
| No copies of e4 | 2,579 (75%) | 2,088 (77%) | 407 (70%) | 84 (64%) |  |
| Any copy of e4 | 861 (25%) | 641 (23%) | 173 (30%) | 47 (36%) |  |
| Missing | 380 | 295 | 71 | 14 |  |

<sup>1</sup>Mean (SD); n (%)
<sup>2</sup>Kruskal-Wallis rank sum test; Fisher's Exact Test for Count Data with simulated p-value (based on 2000 replicates)

Among the primary incident analytic sample who were cognitively normal at baseline, 334 declined to CIND and 63 to dementia during follow up. Those who developed cognitive impairment were more likely to be male, and those declining to dementia were more likely to be *APOE-ε4* carriers (**Table 2**).

**Table 2.** Descriptive statistics for participants included in incident analyses, by incident cognitive status: lasting normal, decline to cognitively impaired non-dementia (CIND), decline to dementia.

| Characteristic | Overall<br>N = 2,703<sup>1</sup> | Lasting<br>normal<br>N = 2,306<sup>1</sup> | Decline to<br>CIND<br>N = 334<sup>1</sup> | Decline to<br>dementia<br>N = 63<sup>1</sup> | p-value<sup>2</sup> |
| --- | --- | --- | --- | --- | --- |
| **Follow up time (years)** |  |  |  |  | <0.001 |
| 2 | 331 (12%) | 190 (8.2%) | 129 (39%) | 12 (19%) |  |
| 4 | 2,372 (88%) | 2,116 (92%) | 205 (61%) | 51 (81%) |  |
| **Age at baseline (2016)** | 69 (9) | 68 (9) | 72 (10) | 74 (9) | <0.001 |
| **Sex** |  |  |  |  | 0.001 |
| Male | 1,089 (40%) | 897 (39%) | 162 (49%) | 30 (48%) |  |
| Female | 1,614 (60%) | 1,409 (61%) | 172 (51%) | 33 (52%) |  |
| **Race/ethnicity** |  |  |  |  | 0.2 |
| NH-White | 1,943 (72%) | 1,677 (73%) | 225 (67%) | 41 (65%) |  |
| NH-Black | 370 (14%) | 303 (13%) | 56 (17%) | 11 (17%) |  |
| NH-Other | 77 (2.8%) | 68 (2.9%) | 8 (2.4%) | 1 (1.6%) |  |
| Hispanic | 313 (12%) | 258 (11%) | 45 (13%) | 10 (16%) |  |
| **Years of education at baseline** | 13.46 (2.75) | 13.67 (2.63) | 12.28 (2.99) | 12.35 (3.80) | <0.001 |
| **Baseline smoke status** |  |  |  |  | 0.035 |
| Never | 1,228 (45%) | 1,069 (46%) | 138 (41%) | 21 (33%) |  |
| Former | 1,188 (44%) | 1,003 (43%) | 149 (45%) | 36 (57%) |  |
| Current | 287 (11%) | 234 (10%) | 47 (14%) | 6 (9.5%) |  |
| **Baseline drink status (#days/week)** | 1.34 (2.07) | 1.37 (2.08) | 1.16 (2.04) | 1.13 (2.03) | 0.029 |
| **Baseline BMI (kg/m2)** | 29.1 (6.3) | 29.2 (6.3) | 29.0 (6.6) | 29.0 (5.7) | 0.8 |
| **Granulocytes %** | 65 (12) | 64 (12) | 65 (13) | 68 (13) | 0.026 |
| **Natural Killer %** | 6.9 (3.7) | 6.8 (3.7) | 7.4 (4.0) | 7.0 (4.0) | 0.031 |
| **B cell %** | 5.2 (3.5) | 5.2 (3.5) | 5.0 (3.4) | 4.7 (3.6) | 0.090 |
| **CD4+ T cell %** | 17 (8) | 17 (8) | 15 (8) | 14 (7) | <0.001 |
| **CD8+ T cell %** | 5.9 (6.7) | 5.7 (6.5) | 6.5 (7.3) | 6.5 (9.2) | 0.3 |
| **Monocytes %** | 7.34 (2.78) | 7.32 (2.78) | 7.51 (2.82) | 7.54 (2.62) | 0.4 |
| **APOE e4 allele carrier status** |  |  |  |  | <0.001 |
| No copies of e4 | 1,876 (77%) | 1,614 (77%) | 233 (77%) | 29 (54%) |  |
| Any copy of e4 | 573 (23%) | 478 (23%) | 70 (23%) | 25 (46%) |  |
| (Missing) | 254 | 214 | 31 | 9 |  |

<sup>1</sup>n (%); Mean (SD)
<sup>2</sup>Fisher's Exact Test for Count Data with simulated p-value (based on 2000 replicates); Kruskal-Wallis rank sum test

### 3.2 Prevalent analysis: single DNA methylation sites associated with cognitive impairment non-dementia

In the primary analyses, we observed 5,322 DNA methylation sites were nominally associated (p-value<0.01) with prevalent CIND versus normal cognition (**Figure 2**). Of these, 56.3% had higher DNA methylation with impairment. The top CpG by p-value was cg00026891 (−0.75 lower percent methylation in CIND group, p-value=4.08x10^-7^) annotated to the gene *KRT16* (distribution of top 10 CpGs shown in **Figure 3**). Summary statistics for all tested CpGs are contained in **Supplemental Table 3** for CIND.

**Figure 2.**
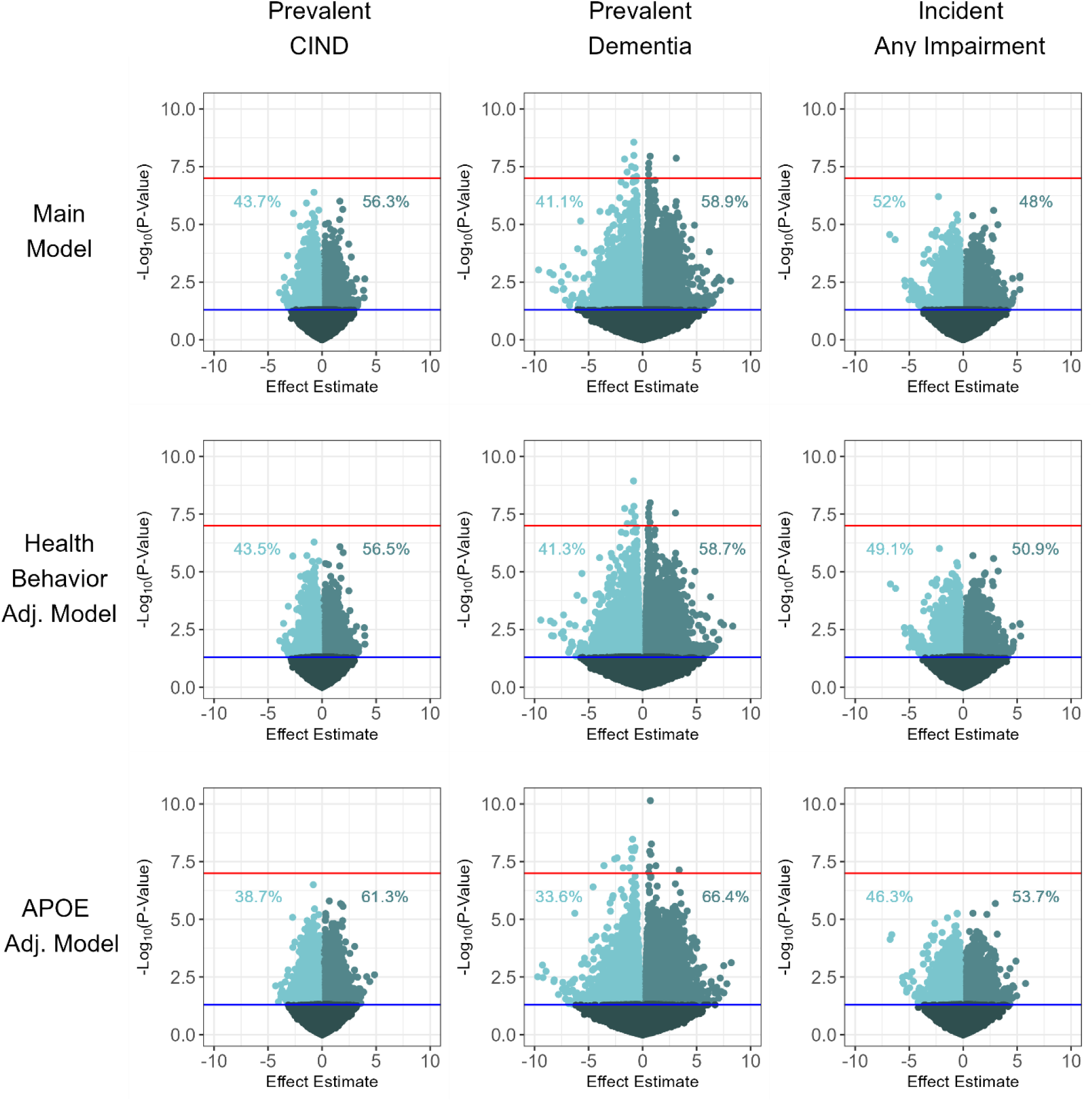
Single CpG site results volcano plots for prevalent cognitively impaired non-dementia (CIND), prevalent dementia, and incident any impairment. Percentage of CpG’s with p-value < 0.01 with increased or decreased methylation are shown.

**Figure 3.**
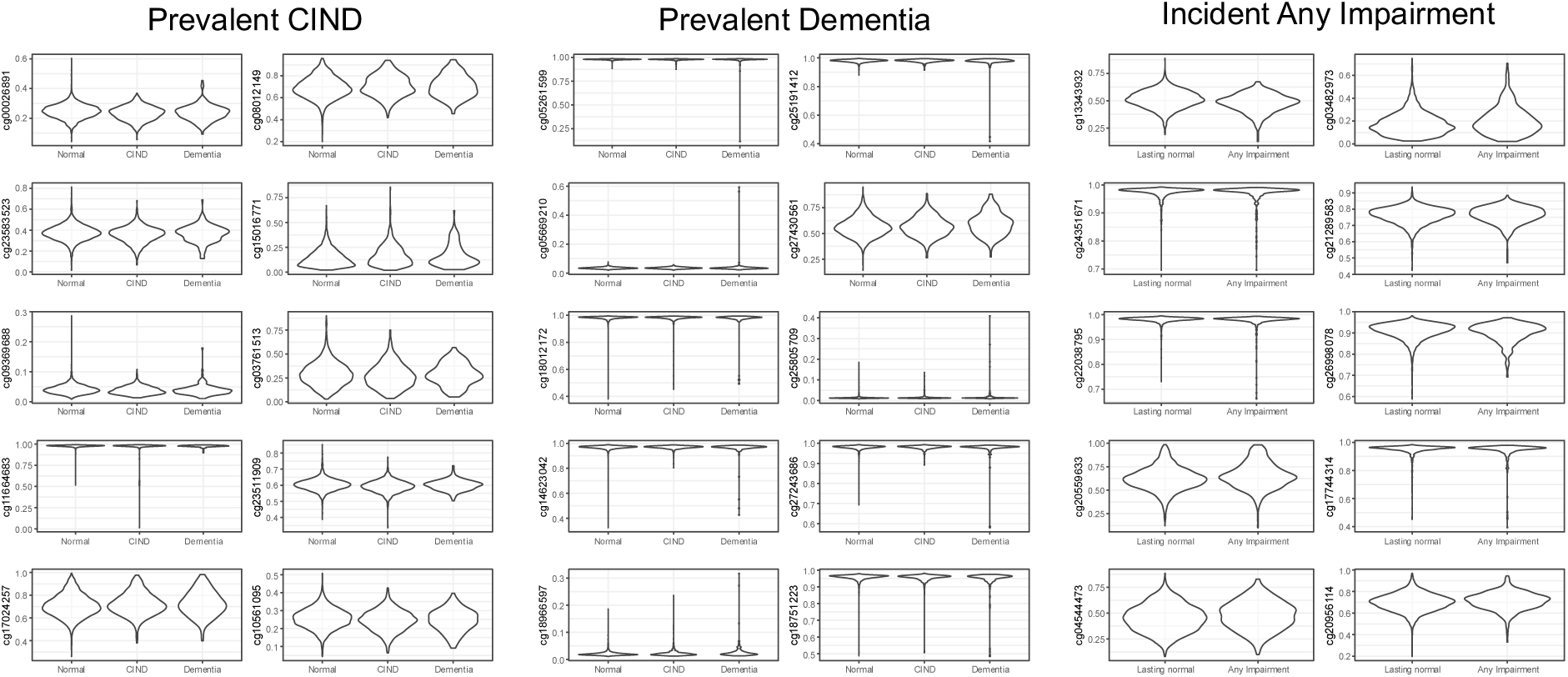
Violin plots showing distribution of DNA methylation values of top 10 CpGs by p-value associated with prevalent cognitively impaired non-dementia (CIND), prevalent dementia, and incident any impairment.

In sensitivity analyses, using the health behavior model we observed 5,264 DNA methylation sites nominally associated with CIND (**Supplemental Table 4**). In the *APOE* sensitivity model, we observed 7,416 DNA methylation sites nominally associated with CIND (**Supplemental Table 5**). A total of 3,509 CpGs overlapped between all three models (primary, health behavior, *APOE*), with the *APOE* model having the most unique top CpGs (n=3,478) (**Supplemental Figure 3**).

There were no or modest levels of genomic inflation for CIND with lambda values of 1.002 for the main model, 0.98 for the health behavior model, and 1.22 for the *APOE* model (**Supplemental Figure 4**).

### 3.3 Prevalent analysis: single DNA methylation sites associated with dementia

In the primary analyses, we observed 14,366 DNA methylation sites nominally associated with prevalent dementia and 58.9% of these sites had higher DNA methylation with dementia (**Figure 2**). For dementia, 166 CpGs met FDR significance (adjusted-p<0.05), and the top CpG was cg05261599 (−0.82 lower percent methylation in dementia, p-value=2.77x10^-9^) (distribution of top 10 CpGs shown in **Figure 3**). Summary statistics for all tested CpGs are contained in **Supplemental Table 6** for dementia.

In the health behavior sensitivity model, we observed 13,954 nominally associated sites (166 FDR significant) (**Supplemental Table 7**). In the *APOE* sensitivity model we observed 14,352 nominally associated sites (129 FDR significant) (**Supplemental Table 8**). Between the main, health behavior, and *APOE* models 9,425 nominal CpGs overlapped between all three models (83 FDR significant in all three models), and the *APOE* model had the most unique nominal CpGs (**Supplemental Figure 3**).

The dementia associations had lambda values of 1.646 for the main model, 1.637 for the health behavior model, and 1.515 for the *APOE* model (**Supplemental Figure 4**).

### 3.4 Incident analysis: single DNA methylation sites associated with cognitive impairment

In the primary incident analysis, we observed the development of any impairment was nominally associated with DNA methylation at 4,898 CpGs, of which 48% had higher methylation (**Figure 2**). The top CpG by p-value was cg13343932 (−2.29 lower percent methylation in any impairment group, p-value=6.32x10^-7^) annotated to the gene *LOC101928674* (distribution of top 10 CpGs shown in **Figure 3**). Summary statistics for all tested CpGs are contained in **Supplemental Table 9**.

In the health behavior sensitivity model, there were 4,997 sites nominally associated with the development of any impairment (**Supplemental Table 10**). In the *APOE* sensitivity model, there were 4,110 sites nominally associated with the development of any impairment (**Supplemental Table 11**). Together, we observed 2,598 of these sites overlapped between all three models (primary, health behavior, *APOE*) (**Supplemental Figure 3**). As in the case with prevalent models, the *APOE* model had the most unique top CpGs (n=1,368).

In an exploratory analysis, we used the three category cognitive variable (CIND, dementia, normal cognition) and ran the primary model (**Supplemental Figure 5**). We observed incident CIND was associated with DNA methylation at 4,208 nominal CpGs (**Supplemental Table 12**). Incident dementia was associated with DNA methylation at 5,792 nominal CpGs (**Supplemental Table 13**). Incident dementia had 256 FDR significant CpGs, and the top CpG was cg20619347 (1.99 higher methylation in dementia, p-value=6.41x10^-15^) annotated to *SNRK*. No FDR significant CpGs were observed for any other analysis.

Lambda values for incident any impairment were 0.983 for the main model, 0.969 for the health behavior model, and 0.858 for the *APOE* model. For incident CIND and incident dementia in the main model, lambdas were 0.873 and 0.885, respectively (**Supplemental Figure 6**).

### 3.5 Biologic pathway enrichment

We tested our cognition-associated DNA methylation sites for enrichment of various factors to interpret patterns in a biologic context. In gene ontology analysis, prevalent CIND CpGs were enriched (adjusted p-value < 0.05) for ligand-gated channel activity and skeletal system development pathways (**Supplementary Table 14**). Dementia CpGs were enriched in pathways related to immune activation, cell movement, structure and development, ion transport, and vesicles (**Supplementary Table 15**). Incident any impairment CpGs were enriched in pathways related to morphogenesis and development, neuron differentiation, and DNA transcription activator activity (**Supplemental Table 16**).

### 3.6 Genomic region enrichment

Compared to the array background, CpGs nominally associated with prevalent CIND had lower proportion in island regions (background 12.1%, CIND 10.9%, chi-square p-value=0.0097), and higher proportion in north shore regions (background 11.2%, CIND 10.3%, chi-square p-value=0.0032) (**Supplemental Figure 7**). CpG island regions for prevalent dementia CpGs did not significantly differ from background of tested CpGs. In incident any impairment analysis, top CpGs were enriched for island regions (background 12.1%, any impairment 18.4%, chi-square p-value<2.2x10^-16^), and depleted in open sea regions (background 60.8%, any impairment 55.5%, chi-square p-value<2.2x10-^16^) (**Supplementary Figure 8**).

### 3.7 Chromatin state enrichment

In chromatin state enrichment for prevalent CIND CpGs, blood cell type signal had highest enrichment, with genic enhancers most enriched (**Supplementary Figure 9**). In prevalent dementia related CpGs, repressed poly comb, bivalent enhancers, bivalent/poised TSS and enhancers signals were generally enriched (**Supplementary Figure 10**). For incident any impairment CpGs, cell type signals were varied, with (weak) repressed polycomb and bivalent enhancer domains enriched (**Supplementary Figure 11**).

### 3.8 Comparison between prevalent and incident findings

In general, prevalent dementia had larger effects estimates and lower p-values compared to prevalent CIND and incident any impairment results (**Figure 2**). For exploratory incident CIND and incident dementia analyses, dementia single site results were similarly larger in effect estimates and significance levels (**Supplemental Figure 5**). Effect estimates for sensitivity models were highly correlated with main model estimates. Health behavior model effect estimates had almost 1.00 correlation with main model estimates for prevalent and incident models. The *APOE* models were also highly correlated with main models, with Pearson correlation of 0.93 in prevalent analysis, and 0.92 in incident analysis (**Supplemental Figure 12**).

Effect estimates were in the same direction across all three conditions, except for cg08648606, which had lower methylation for incident any impairment (p-value<0.01), and higher methylation for prevalent CIND (p-value<0.01) and dementia (p-value=0.1). Between the prevalent CIND, prevalent dementia and incident any impairment analyses, 9 CpGs were nominally associated in all 3 analyses (**Supplementary Figure 13**). These CpGs included cg02583484 (annotated to *HNRNPA1*), cg15266133 (annotated to *LOC102724084*), cg24287460 (annotated to *CCDC48*), cg17124509 (annotated to *C17orf57*), and cg02553054 (annotated to *SMARCD1*) (**Table 3**).

**Table 3.** Top CpGs (p-value < 0.01) overlapping between prevalent cognitively impaired non-dementia (CIND), prevalent dementia, and incident any cognitive impairment analyses.

| CpG | Gene | Prevalent CIND Estimate | Prevalent CIND P | Prevalent Dementia Estimate | Prevalent Dementia P | Incident Any Impairment Estimate | Incident Any Impairment P |
| --- | --- | --- | --- | --- | --- | --- | --- |
| cg02583484 | HNRNPA1 | -0.8009 | 0.00057 | -1.366 | 0.00265 | -0.7299 | 0.00994 |
| cg25144382 |  | 0.88 | 0.00163 | 1.675 | 0.00217 | 1.278 | 0.000223 |
| cg08648606 |  | 0.2055 | 0.00278 | 0.3555 | 0.00813 | -0.2204 | 0.00953 |
| cg15266133 | LOC102724084 | 0.3475 | 0.00811 | 0.7298 | 0.00445 | 0.4741 | 0.00504 |
| cg24287460 | CCDC48 | 0.3702 | 0.00841 | 0.7084 | 0.00988 | 0.461 | 0.00703 |
| cg04885881 |  | -0.8399 | 0.00901 | -2.032 | 0.00124 | -1.397 | 0.000473 |
| cg17124509 | C17orf57 | 0.4757 | 0.00938 | 1.027 | 0.00414 | 0.5609 | 0.00582 |
| cg07974619 |  | 0.7046 | 0.00941 | 1.37 | 0.00982 | 0.9863 | 0.00362 |
| cg02553054 | SMARCD1 | -0.534 | 0.00942 | -1.169 | 0.00363 | -1.046 | 5.50x10<sup>-5</sup> |

### 3.9 Replication testing results

There was minimal overlap with CpGs previously identified in peripheral blood associated with Alzheimer’s disease or episodic memory. We used X criteria to test for overlap. CpGs associated with prevalent dementia in HRS had most overlap with Roubroeks 2020 CpGs: 13 overlapping with mild cognitive impairment, and 24 with Alzheimer’s disease (**Supplemental Figure 13**). Similarly, in terms of overlap with CpGs associated with episodic memory in Sommerer 2022, prevalent dementia had most overlap with cross-sectional (120 sites) and longitudinal (107 sites) episodic memory (**Supplemental Figure 14**).

## 4 DISCUSSION

Our study sought to better understand DNA methylation differences among a large population of older adults who either go on to develop cognitive impairment or dementia, or who maintain normal cognition during follow up. In a large sample of older and demographically diverse US adults from the HRS, we assessed blood-based DNA methylation for associations with prevalent (n=3,395) and incident (n=2,401) cognitive status. We observed many single CpG sites to have nominal associations (p<0.01) with our primary outcomes: 5,322 CpGs observed for prevalent CIND, 14,366 CpGs observed for prevalent dementia, 4,898 CpGs observed for incident any cognitive impairment. Among our primary outcomes the top CpG sites by p-value were cg00026891 for prevalent CIND, cg05261599 for prevalent dementia, and cg13343932 for incident any impairment. After correcting for multiple comparisons, 166 CpGs were significantly associated with prevalent dementia, and 256 CpGs were significantly associated with incident dementia. Nine CpGs overlapped all analyses including cg02583484 (*HNRNPA1*), cg15266133 (*LOC102724084*), cg24287460 (*CCDC48*), cg17124509 (*C17orf57*), and cg02553054 (*SMARCD1*). We performed multiple sensitivity analyses including models for health behavior and *APOE-ε4* carrier status to provide greater rigor. Together, our findings suggest DNA methylation measured in blood may be early markers of cognitive impairment.

Chief among our observed prevalent CIND associations was site cg00026891, annotated to the *KRT16* gene, and which encodes the protein keratin 16. Molecular physiologic processes regulated by keratin 16 functions not typically associated with keratin genes were identified such as regulation of cell growth, migration, proliferation, apoptosis, and response to DNA damage.^37^ A number of studies have identified upregulation of keratin 16 in roles related to carcinogenesis, regulation of mitochondrial morphology, and oxidative stress.^37–40^ In addition, *KRT16* gene expression may be involved in Alzheimer’s disease pathogenesis via the Wnt receptor signaling pathway.^41^ Growing evidence supports the role of Wnt signaling in Alzheimer’s disease.^41–43^ Inhibition of normal signaling allows for beta-amyloid mediated synaptic loss, and increased Wnt signaling could be protective for brain synapses in patients with Alzheimer’s disease.^41^ Taken together, upregulation of *KRT16* may influence Wnt signaling and could contribute to dementia pathology. Thus, the novel results of this study suggest new potential avenues to explore in future studies of DNA methylation and dementia research of blood-based biomarkers.

Among our top CpG hits for our exploratory incident dementia analysis was cg20619347, annotated to the sucrose nonfermenting 1-related kinase (*SNRK*) gene. *SNRK* is a member of the adenosine monophosphate activated protein kinase (*AMPK*) family, and is a key mediator in cellular metabolic homeostasis.^44^ Within the neuronal system, *SNRK* regulates low extracellular potassium induced apoptosis in cerebral neurons.^44^ *SNRK* also plays a role in inflammatory pathways and insulin resistance and has been suggested as a target for treating type 2 diabetes.^45^ Neuronal cell death, inflammation, and altered energy homeostasis are well established hallmarks of neurodegenerative diseases.^46^ To date few studies have noted *SNRK* among top hits in blood based DNA methylation associations for dementia. Since *SNRK* participates in these processes, it may be an important candidate for future dementia blood-based biomarker panels.

We observed the *HNRNPA1* gene to have associations with our three main models of prevalent CIND, prevalent dementia, and incident any cognitive impairment. Heterogeneous nuclear ribonucleoproteins (*hnRNPs*) a family of RNA binding proteins are integral for RNA metabolism with functions such as stabilization, and translational regulation.^47^ *HNRNPA1* is associated with dysregulated RNA metabolism, an important mechanism underpinning pathogenesis of frontotemporal dementia and ALS in previous studies.^48,49^ To date this is the first study to connect *HNRNPA1* gene to prevalent and incident cognitive impairment.

Our study showed DNA methylation sites associated with cognition were enriched in genes associated with ion transport and ligand-gated channel activity. Two of our top hits (*CCDC48, C17orf57*) associated with our three main models of prevalent CIND, prevalent dementia, and incident any cognitive impairment have calcium ion functionality. *CCDC48*, also referred to as *EFCC1,* is predicted to enable calcium ion binding activity.^50^ Calcium ion binding activity is crucial for proper brain functioning, as it regulates key functions such as synaptic transmission and cell survival.^51^ Dysregulated calcium signaling has been observed in various neurodegenerative diseases such as Alzheimer’s and Parkinson’s.^51^ *C17orf57 (*synonym: *EFCAB13*), an EF-handed calcium binding domain, has been observed to increase risk for Alzheimer’s disease in a study by Manikandan et al.^52,53^ Studies have observed low calcium ion binding after phosphorylation of the *EFhd2 gene* to interact with tau. ^54^ Overall, calcium ion binding may underpin key pathways for dementia with this study novelly suggesting *CCDC48 and C17orf57* genes as future areas of study.

DNA methylation sites associated with cognition were further enriched in gene ontology functions of morphology and development. The *SMARCD1* gene was also observed to have nominal associations with prevalent CIND, prevalent dementia, and incident any impairment analyses. *SMARCD1* is a member of the ATP-dependent chromatin remodeling complex SWI/SNF, has key roles in transcription regulation, and gene expression regulation.^55,56^ Neuronal gene expression regulation is crucial for both brain development and normal cognitive function in adulthood.^57^ The SWI/SNF complex is also important for synaptic plasticity and memory in mature neurons.^57,58^ The role of *SMARCD1* in brain development points to a potential link to age-related neurological function, as the brain is sensitive to change at any stage of life, dysregulation of neural development could set the stage for neurological diseases like dementia in adulthood.^56,57,59^ Potentially, disruptions in chromatin remodeling could contribute to the pathology of dementia, as it pathologically contributes to neurodevelopmental disorders and some cancers.^56–59^ While *SMARCD1* has not been linked to dementia, it can been tied to plaque progression in atherosclerosis, a risk factor for vascular dementia.^60,61^ Taken together, *SMARCD1* may be a promising biomarker for dementia and cognitive impairment.

While few of our top hits have been discussed in the current literature as biomarker candidates for dementia, many of them have been highlighted in epigenome-wide association studies of related diseases such as atherosclerosis and diabetes.^45,60,61^ In addition, we have identified genes that are involved with signaling pathways connected to Alzheimer’s disease pathology. DNA methylation at identified sites are potentially early identifiers for dementia on the disease continuum, or markers of important risk factors.

### 4.1 Limitations and future directions

Limitations of this study include a small sample size of incident dementia which led us to combine CIND and dementia into one category of any cognitive impairment. Other studies with a greater period of longitudinal dementia data, or in the future with more years of follow up in the HRS, may be more well powered to examine incident dementia. We observed modest overlap with CpG’s previously identified in peripheral blood associated with episodic memory or Alzheimer’s Disease. However, this is in line with replication results reported by other studies.^35,62^ One explanation could be that episodic memory, Alzheimer’s Disease, and dementia are not necessarily equivalent, and if the observed EWAS associations were related outcome specific risk factors, the set of risk factors in each study population could be different. Additionally, limited availability of CpG sites to perform replication, and differences in the population demographics of each study may contribute to the minimal replication.

One CpG site cg08648606 meet significance levels (p-value<0.05) for two of our main phenotypes (incident any impairment, prevalent CIND), but had lower methylation for incident any impairment, and higher methylation for prevalent CIND and dementia. Potentially, the heterogeneity between dementia subtypes could contribute to this observation. Subtle differences in circulating cell types between those with dementia vs. cognitive impairment non-dementia have been documented.^35,36,63^ Combining incident dementia and incident CIND into a general incident any cognitive impairment outcome to increase sample size may have impaired our ability to detect association signals. The long prodromal stage of dementia may lead to discordant diagnosis vs. disease status, where participants may not have been diagnosed yet but met the clinical criteria for diagnosis, which may mask association signals between DNAm and incident impairment.

While we demonstrated the robustness of our model results, blood-based measures are only a proxy for changes occurring in the brain. To fully understand the highly complex multifactorial etiology of dementia incorporating neuroimaging and neuropathology data, as well as exposures from the environment in childhood and mid-life are important. Future studies incorporating the aforementioned characteristics are paramount to further characterization of preclinical epigenetic changes and successful identification of at-risk populations.^6,16^

### 4.2 Strengths

The longitudinal study design and participant population with diverse demographics and genetic ancestries are key strengths of this analysis. In addition, our large analytic sample size affords this study adequate statistical power to interrogate associations between DNA methylation and cognitive impairment, to help establish DNA methylation signatures with a temporal relationship with the dementia disease continuum. The high correlation of effect estimates between sensitivity models and main models indicate robustness of the models to appropriately identify associations and strengthen the confidence of reported results. Including intermediate dementia disease state of CIND provides insight into how DNA methylation may differ at different stages of cognitive decline.

### 4.3 Summary and implications

Overall, this study offers significant contributions to the rapidly growing research on blood-based biomarkers of dementia by identifying novel genes associated with both incident and prevalent cognitive impairment, in a robust analysis. Our results provide promising grounds for non-invasive blood based early detection of cognitive impairment and future studies focused on replication and validation of findings are needed. Importantly, we address common gaps in the literature by including a large sample size, longitudinal component, and diverse demographic and genetic participant profiles.

## Data Availability

All demographic and cognitive data used in this analysis are publicly available and can be downloaded from the Health and Retirement Study. DNA methylation data is restricted. Access requires completing a restricted data agreement with the HRS. R code for statistical analysis is available on GitHub.

https://hrs.isr.umich.edu/

https://github.com/bakulskilab/HRS-DNAm-cognition

## Acknowledgements

We appreciate the participants and staff of the Health and Retirement Study.

## Consent statement

All study participants provided written informed consent.

## Data availability statement

All demographic and cognitive data used in this analysis are publicly available and can be downloaded from the Health and Retirement Study at https://hrs.isr.umich.edu/. DNA methylation data is restricted. Access requires completing a restricted data agreement with the HRS. R code for statistical analysis is available on GitHub (https://github.com/bakulskilab/HRS-DNAm-cognition).

