## Supplemental files for "DNA methylation-wide association study of prevalent and incident dementia in the US Health and Retirement Study"

15 Supplementary Figures

16 Supplementary Tables

**Supplemental Figure 1.** DNA methylation quality control summary. Samples (left) and probes (right) were dropped based on quality control criteria described.

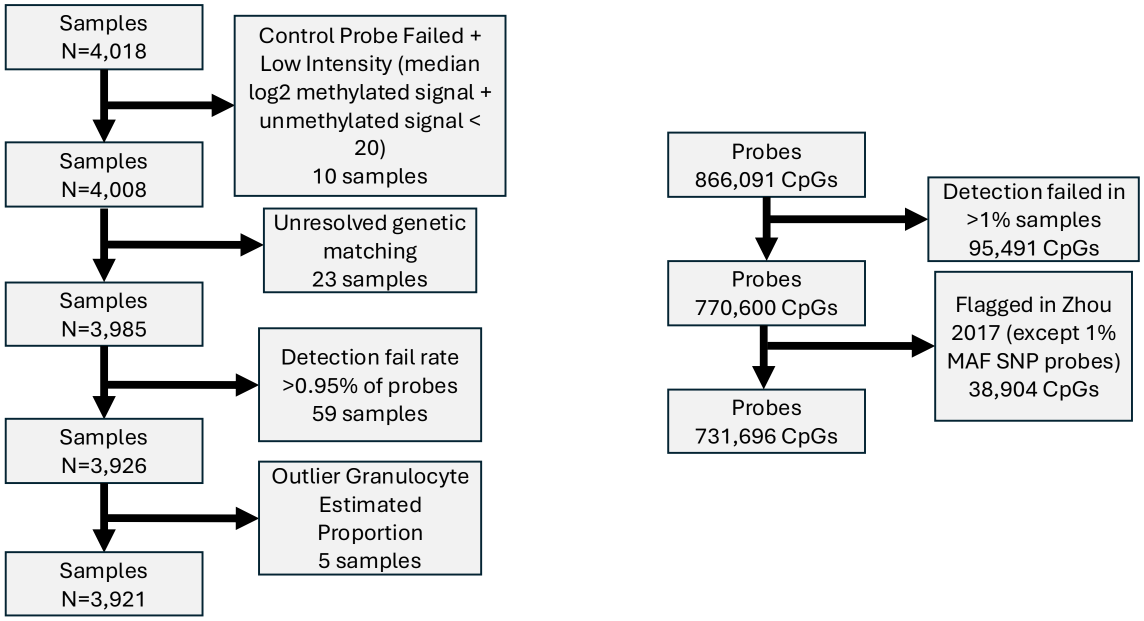

**Supplemental Figure 2.** Flowchart for inclusion and exclusion in A.) cross-sectional analysis, and B.) incident analysis.

**A.**

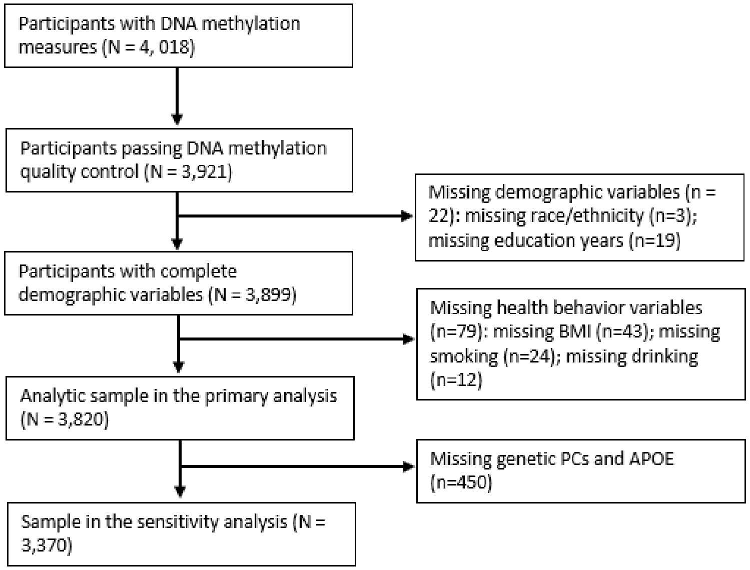


**B.**

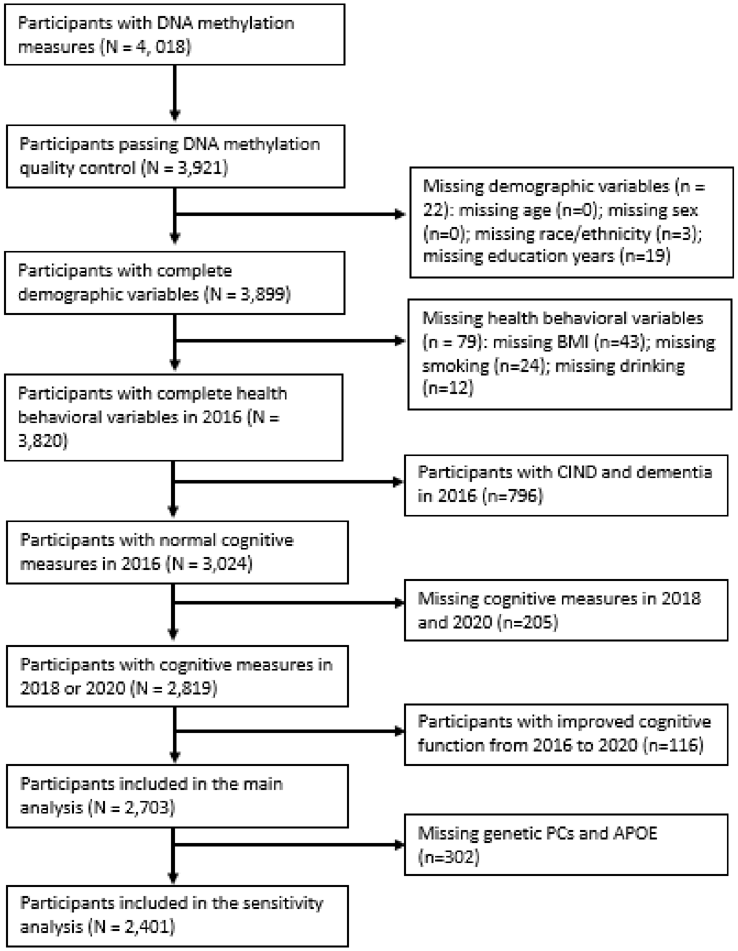

**Supplemental Table 1**. Excluded and included sample for cross-sectional analyses.

| Characteristic | Overall<br>N = 4,018[1] | Excluded<br>N = 198[1] | Included<br>N = 3,820[1] | p-value[2] |
|---|---|---|---|---|
| **Langa-Weir classification 2016** | | | | >0.9 |
| Normal | 3,183 (79%) | 159 (80%) | 3,024 (79%) | |
| CIND | 683 (17%) | 32 (16%) | 651 (17%) | |
| Dementia | 152 (3.8%) | 7 (3.5%) | 145 (3.8%) | |
| **Age at baseline (2016)** | 70 (10) | 68 (10) | 70 (10) | <0.001 |
| **Sex** | | | | 0.4 |
| Male | 1,669 (42%) | 76 (38%) | 1,593 (42%) | |
| Female | 2,349 (58%) | 122 (62%) | 2,227 (58%) | |
| **Race/ethnicity** | | | | 0.047 |
| NH-White | 2,669 (66%) | 115 (59%) | 2,554 (67%) | |
| NH-Black | 657 (16%) | 37 (19%) | 620 (16%) | |
| NH-Other | 122 (3.0%) | 4 (2.1%) | 118 (3.1%) | |
| Hispanic | 567 (14%) | 39 (20%) | 528 (14%) | |
| (Missing) | 3 | 3 | 0 | |
| **Years of education at baseline** | 12.8 (3.2) | 12.4 (3.7) | 12.8 (3.2) | 0.2 |
| (Missing) | 19 | 19 | 0 | |
| **Baseline smoke status** | | | | 0.086 |
| Never | 1,764 (44%) | 79 (45%) | 1,685 (44%) | |
| Former | 1,775 (44%) | 67 (39%) | 1,708 (45%) | |
| Current | 455 (11%) | 28 (16%) | 427 (11%) | |
| (Missing) | 24 | 24 | 0 | |
| **Baseline drink status (#days/week)** | 1.19 (2.01) | 1.08 (1.90) | 1.20 (2.01) | 0.4 |
| (Missing) | 12 | 12 | 0 | |
| **Baseline BMI (kg/m2)** | 28.9 (6.3) | 28.5 (5.7) | 28.9 (6.3) | 0.6 |
| (Missing) | 43 | 43 | 0 | |
| **Granulocytes percent - %** | 65 (13) | 64 (12) | 65 (13) | >0.9 |
| **Natural Killer cell percent - %** | 7.1 (3.8) | 6.9 (3.8) | 7.1 (3.8) | 0.7 |
| **B cell %** | 5.2 (3.8) | 5.6 (3.5) | 5.2 (3.8) | 0.095 |
| **CD4+ T cell %** | 16 (8) | 17 (9) | 16 (8) | 0.6 |

| **Characteristic** | **Overall**<br>N = 4,018[1] | **Excluded**<br>N = 198[1] | **Included**<br>N = 3,820[1] | **p-value**[2] |
|---|---|---|---|---|
| **CD8+ T cell %** | 6.1 (6.8) | 5.7 (6.5) | 6.1 (6.8) | 0.8 |
| **Monocytes %** | 7.41 (2.90) | 6.77 (2.99) | 7.42 (2.90) | 0.012 |
| **APOE e4 allele carrier status** | | | | 0.5 |
| No copies of e4 | 2,695 (75%) | 116 (72%) | 2,579 (75%) | |
| Any copy of e4 | 905 (25%) | 44 (28%) | 861 (25%) | |
| (Missing) | 418 | 38 | 380 | |

[1]n (%); Mean (SD)

[2]Fisher's exact test; Wilcoxon rank sum test

**Supplementary Table 2.** Excluded and included sample for incident analyses.

| Characteristic | Overall<br>N = 4,018[1] | Excluded<br>N = 1,315[1] | Included<br>N = 2,703[1] | p-value[2] |
|---|---|---|---|---|
| **Langa-Weir classification 2016** | | | | <0.001 |
| Normal | 3,183 (79%) | 480 (37%) | 2,703 (100%) | |
| CIND | 683 (17%) | 683 (52%) | 0 (0%) | |
| Dementia | 152 (3.8%) | 152 (12%) | 0 (0%) | |
| **Langa-Weir classification 2018** | | | | <0.001 |
| Normal | 2,835 (81%) | 400 (44%) | 2,435 (94%) | |
| CIND | 528 (15%) | 372 (41%) | 156 (6.0%) | |
| Dementia | 153 (4.4%) | 141 (15%) | 12 (0.5%) | |
| (Missing) | 502 | 402 | 100 | |
| **Langa-Weir classification 2020** | | | | <0.001 |
| Normal | 2,544 (78%) | 428 (55%) | 2,116 (86%) | |
| CIND | 505 (16%) | 217 (28%) | 288 (12%) | |
| Dementia | 194 (6.0%) | 138 (18%) | 56 (2.3%) | |
| (Missing) | 775 | 532 | 243 | |
| **Follow up cognitive function** | | | | 0.001 |
| Lasting normal | 2,439 (86%) | 133 (96%) | 2,306 (85%) | |
| Decline to CIND | 339 (12%) | 5 (3.6%) | 334 (12%) | |
| Decline to dementia | 64 (2.3%) | 1 (0.7%) | 63 (2.3%) | |
| (Missing) | 1,176 | 1,176 | 0 | |
| **Follow up time (years)** | | | | 0.8 |
| 2 | 349 (12%) | 18 (13%) | 331 (12%) | |
| 4 | 2,493 (88%) | 121 (87%) | 2,372 (88%) | |
| (Missing) | 1,176 | 1,176 | 0 | |
| **Age at baseline (2016)** | 70 (10) | 72 (11) | 69 (9) | <0.001 |
| **Sex** | | | | 0.022 |
| Male | 1,669 (42%) | 580 (44%) | 1,089 (40%) | |
| Female | 2,349 (58%) | 735 (56%) | 1,614 (60%) | |
| **Race/ethnicity** | | | | <0.001 |
| NH-White | 2,669 (66%) | 726 (55%) | 1,943 (72%) | |
| NH-Black | 657 (16%) | 287 (22%) | 370 (14%) | |

| Characteristic | Overall<br>N = 4,018[1] | Excluded<br>N = 1,315[1] | Included<br>N = 2,703[1] | p-value[2] |
|---|---|---|---|---|
| NH-Other | 122 (3.0%) | 45 (3.4%) | 77 (2.8%) | |
| Hispanic | 567 (14%) | 254 (19%) | 313 (12%) | |
| (Missing) | 3 | 3 | 0 | |
| **Years of education at baseline** | 12.8 (3.2) | 11.5 (3.7) | 13.5 (2.8) | <0.001 |
| (Missing) | 19 | 19 | 0 | |
| **Baseline smoke status** | | | | 0.024 |
| Never | 1,764 (44%) | 536 (42%) | 1,228 (45%) | |
| Former | 1,775 (44%) | 587 (45%) | 1,188 (44%) | |
| Current | 455 (11%) | 168 (13%) | 287 (11%) | |
| (Missing) | 24 | 24 | 0 | |
| **Baseline drink status (#days/week)** | 1.19 (2.01) | 0.89 (1.83) | 1.34 (2.07) | <0.001 |
| (Missing) | 12 | 12 | 0 | |
| **Baseline BMI (kg/m2)** | 28.9 (6.3) | 28.4 (6.2) | 29.1 (6.3) | <0.001 |
| (Missing) | 43 | 43 | 0 | |
| **Granulocytes %** | 65 (13) | 65 (14) | 65 (12) | 0.6 |
| **Natural Killer cell %** | 7.1 (3.8) | 7.5 (4.0) | 6.9 (3.7) | <0.001 |
| **B cell %** | 5.2 (3.8) | 5.3 (4.4) | 5.2 (3.5) | 0.7 |
| **CD4+ T cell %** | 16 (8) | 15 (8) | 17 (8) | <0.001 |
| **CD8+ T cell %** | 6.1 (6.8) | 6.5 (7.0) | 5.9 (6.7) | 0.008 |
| **Monocytes %** | 7.41 (2.90) | 7.54 (3.15) | 7.34 (2.78) | 0.2 |
| **APOE e4 allele carrier status** | | | | <0.001 |
| No copies of e4 | 2,695 (75%) | 819 (71%) | 1,876 (77%) | |
| Any copy of e4 | 905 (25%) | 332 (29%) | 573 (23%) | |
| (Missing) | 418 | 164 | 254 | |

[1]n (%); Mean (SD)

[2]Fisher's Exact Test for Count Data with simulated p-value (based on 2000 replicates); Fisher's Exact Test for Count Data; Wilcoxon rank sum test

**Supplemental Table 3**. CpG site results for prevalent CIND analysis.

File: Supplemental Table 3 Prevalent CIND Single Site Results.csv

(top 10 below)

| CpG | Chr | Pos | Gene | CpG Island Region | Effect Estimate | SE | P-Value | Adj. P-Value | Average DNAm |
|---|---|---|---|---|---|---|---|---|---|
| cg00026891 | chr17 | 39769294 | KRT16 | OpenSea | -0.7452 | 0.1469 | 4.08E-07 | 0.23 | 24.39 |
| cg08012149 | chr6 | 45503747 | RUNX2 | OpenSea | 1.653 | 0.3372 | 9.96E-07 | 0.23 | 69.51 |
| cg23583523 | chr11 | 1.18E+08 | AMICA1 | OpenSea | -1.447 | 0.2974 | 1.20E-06 | 0.23 | 37.01 |
| cg15016771 | chr2 | 2.35E+08 | ARL4C | N_Shore | 1.912 | 0.4037 | 2.25E-06 | 0.28 | 14.98 |
| cg09369688 | chr7 | 1.51E+08 | FASTK | N_Shore | -0.3103 | 0.0657 | 2.40E-06 | 0.28 | 4.08 |
| cg03761513 | chr6 | 1.69E+08 | | N_Shore | -2.644 | 0.568 | 3.36E-06 | 0.29 | 28.77 |
| cg11664683 | chr12 | 1.13E+08 | | OpenSea | -0.8932 | 0.1921 | 3.43E-06 | 0.29 | 97.34 |
| cg23511909 | chr3 | 1.28E+08 | RPN1 | S_Shelf | -0.7013 | 0.1533 | 4.94E-06 | 0.36 | 60.15 |
| cg17024257 | chr3 | 1.72E+08 | PLD1 | S_Shore | 1.625 | 0.3589 | 6.19E-06 | 0.4 | 71.04 |
| cg10561095 | chr22 | 50966110 | SCO2 | Island | -0.6243 | 0.1397 | 8.09E-06 | 0.42 | 25.4 |

**Supplemental Table 4**. CpG site results for prevalent CIND analysis health behavior sensitivity model.

File: Supplemental Table 4 Prevalent CIND Health Behavior Sens Single Site Results.csv

**Supplemental Table 5** CpG site results for prevalent CIND analysis APOE sensitivity model.

File: Supplemental Table 5 Prevalent CIND APOE Sens Single Site Results.csv

**Supplemental Figure 3.** Upset plots showing overlap of top CpGs (p-value<0.01) for main, health behavior, and APOE models for prevalent cognitively impaired non-dementia (CIND), prevalent dementia, and incident any impairment analyses.

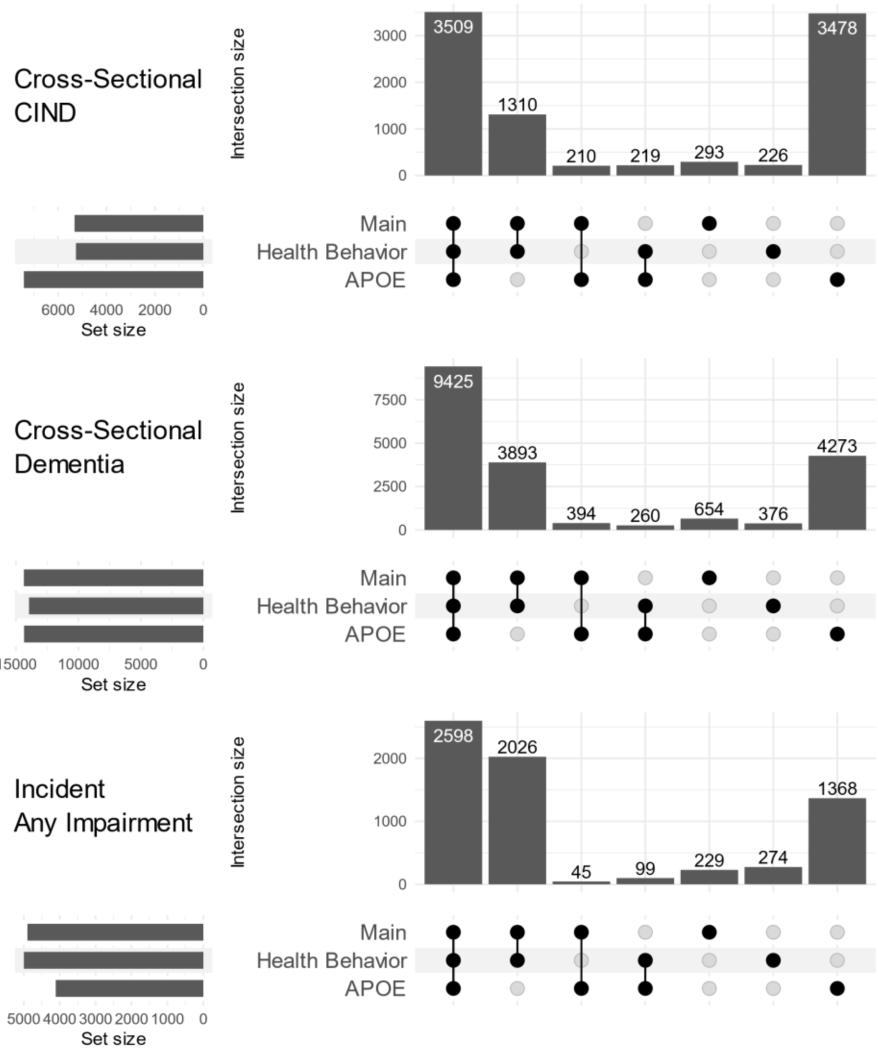

**Supplemental Figure 4**. Model diagnostic QQ-plots for cross-sectional CIND and dementia models.

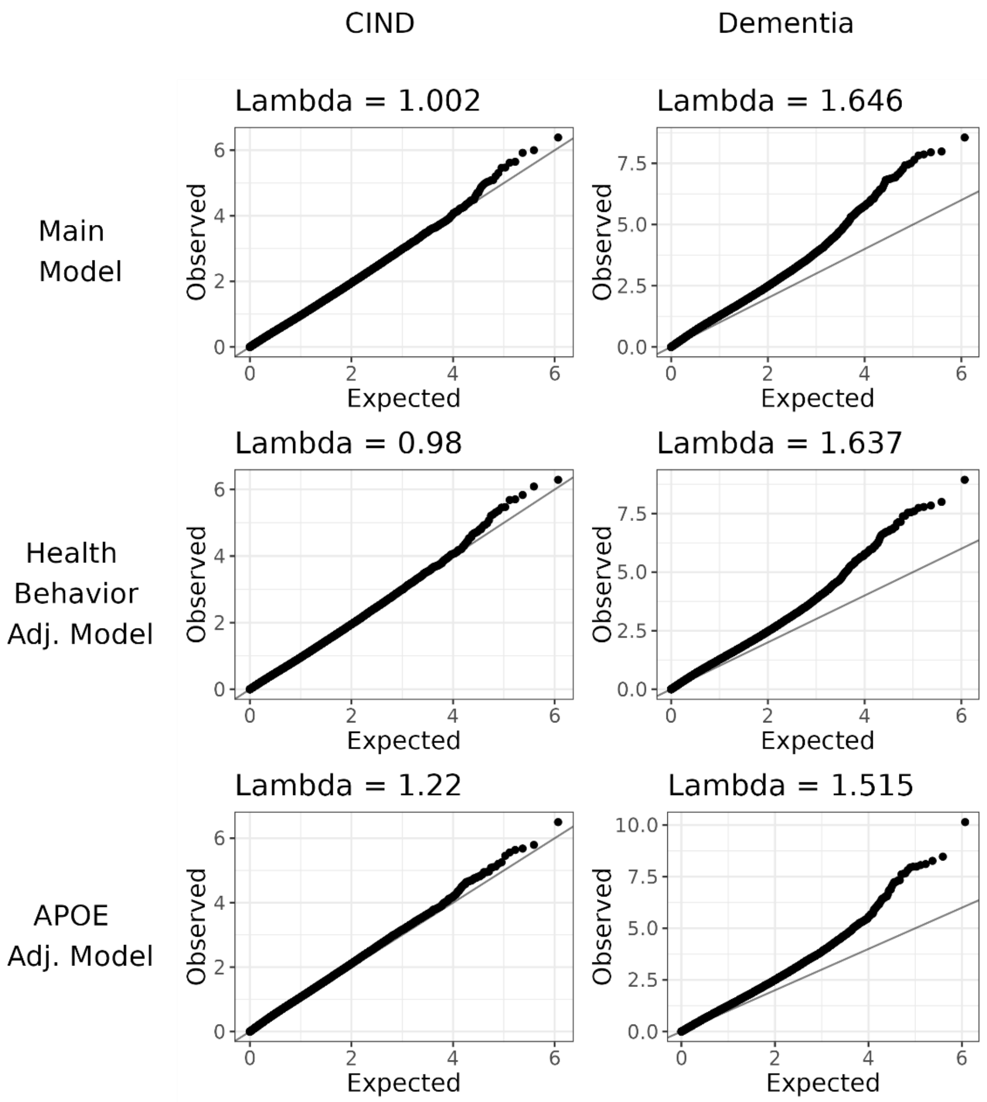

**Supplemental Table 6**. CpG site results for prevalent dementia analysis.

File: Supplemental Table 6 Prevalent Dementia Single Site Results.csv

(top 10 below)

| CpG | Chr | Pos | Gene | CpG Island Region | Effect Estimate | SE | P-Value | Adj. P-Value | Average DNAm |
|---|---|---|---|---|---|---|---|---|---|
| cg05261599 | chr2 | 2.41E+08 | | OpenSea | -0.8161 | 0.137 | 2.77E-09 | 0.0016 | 97.84 |
| cg25191412 | chr16 | 2948694 | FLYWCH2 | OpenSea | -0.8112 | 0.1414 | 1.03E-08 | 0.0017 | 97.95 |
| cg05669210 | chr7 | 1E+08 | SLC12A9 | S_Shore | 0.7008 | 0.1224 | 1.11E-08 | 0.0017 | 3.57 |
| cg27430561 | chr7 | 23633208 | | N_Shelf | 3.113 | 0.547 | 1.36E-08 | 0.0017 | 56.4 |
| cg18012172 | chr16 | 3271087 | | OpenSea | -1.672 | 0.2946 | 1.49E-08 | 0.0017 | 97.92 |
| cg25805709 | chr3 | 44596770 | ZNF167 | Island | 0.581 | 0.1037 | 2.25E-08 | 0.0022 | 1.41 |
| cg14623042 | chr14 | 1.03E+08 | TRAF3 | OpenSea | -1.016 | 0.1832 | 3.13E-08 | 0.0024 | 96.81 |
| cg27243686 | chr12 | 9478612 | | OpenSea | -0.6888 | 0.1248 | 3.59E-08 | 0.0024 | 97.94 |
| cg18966597 | chr7 | 1.55E+08 | | Island | 0.5459 | 0.0990 1 | 3.75E-08 | 0.0024 | 2.15 |
| cg18751223 | chr19 | 1480049 | C19orf25 | N_Shore | -1.43 | 0.2625 | 5.43E-08 | 0.0032 | 95.56 |

**Supplemental Table 7**. CpG site results for prevalent dementia analysis health behavior sensitivity model.

File: Supplemental Table 7 Prevalent Dementia Health Behavior Sens Single Site Results.csv

**Supplemental Table 8**. CpG site results for prevalent dementia analysis APOE sensitivity model.

File: Supplemental Table 8 Prevalent Dementia APOE Sens Single Site Results.csv

**Supplemental Table 9**. CpG site results for incident any impairment analysis.

File: Supplemental Table 9 Incident Any Impairment Single Site Results.csv

(top 10 below)

| CpG | Chr | Pos | Gene | CpG Island Region | Effect Estimate | SE | P-Value | Adj. P-Value | Average DNAm |
|---|---|---|---|---|---|---|---|---|---|
| cg13343932 | chr17 | 76355061 | LOC101928674 | Island | -2.286 | 0.4579 | 6.32E-07 | 0.37 | 50.79 |
| cg03482973 | chr13 | 112756120 | | N_Shelf | 2.824 | 0.5983 | 2.48E-06 | 0.61 | 18.44 |
| cg24351671 | chr18 | 47844186 | | OpenSea | -0.5818 | 0.1255 | 3.74E-06 | 0.61 | 97.29 |
| cg21289583 | chr14 | 100786808 | | OpenSea | 0.8844 | 0.1918 | 4.19E-06 | 0.61 | 76.14 |
| cg22038795 | chr12 | 125920131 | TMEM132B | OpenSea | -0.504 | 0.1117 | 6.67E-06 | 0.72 | 97.86 |
| cg26998078 | chr16 | 815169 | MSLN | OpenSea | -0.9695 | 0.2194 | 1.03E-05 | 0.72 | 90.71 |
| cg20559633 | chr8 | 142734242 | | Island | 3.218 | 0.7285 | 1.04E-05 | 0.72 | 62.79 |
| cg17744314 | chr10 | 89422524 | PAPSS2 | S_Shelf | -1.035 | 0.2362 | 1.23E-05 | 0.72 | 94.75 |
| cg04544473 | chr4 | 153021978 | LOC100996286 | OpenSea | 2.024 | 0.4656 | 1.44E-05 | 0.72 | 45.69 |
| cg20956114 | chr7 | 157451363 | PTPRN2 | N_Shelf | 1.95 | 0.4506 | 1.56E-05 | 0.72 | 70.03 |

**Supplemental Table 10**. CpG site results for incident any impairment analysis health behavior sensitivity model.

File: Supplemental Table 10 Incident Any Impairment Health Behavior Sensitivity Single Site Results.csv

**Supplemental Table 11.** CpG site results for incident any impairment analysis APOE sensitivity model

File: Supplemental Table 11 Incident Any Impairment APOE Sensitivity Single Site Results.csv

**Supplemental Figure 5.** Single CpG site results volcano plots for incident sensitivity analysis, separating CIND and dementia. Percentage of CpG's with p-value < 0.01 with increased or decreased methylation are shown.

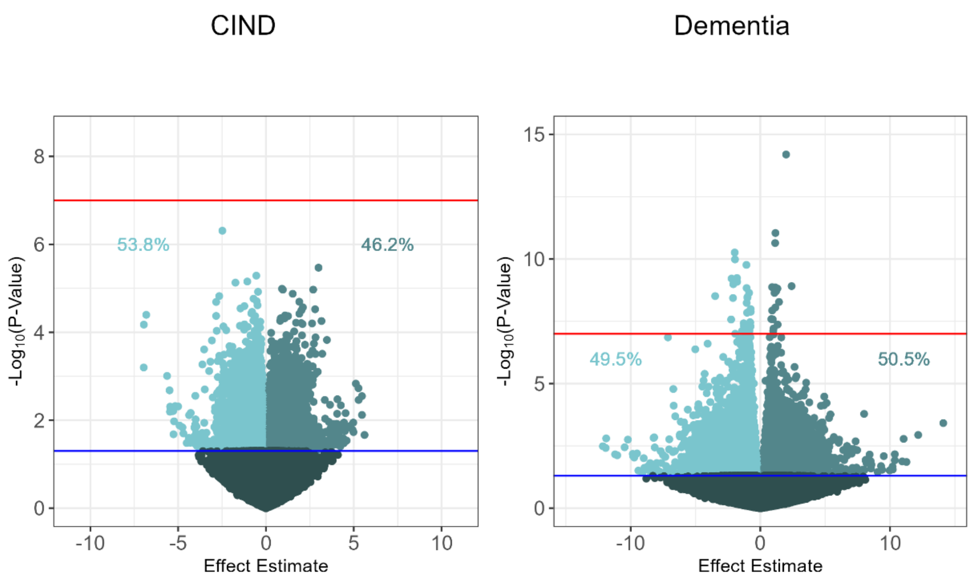


**Supplemental Table 12.** CpG site results for incident CIND sensitivity analysis.

File: Supplemental Table 12 Incident CIND Single Site Results.csv

**Supplemental Table 13**. CpG site results for incident dementia sensitivity analysis.

File: Supplemental Table 13 Incident Dementia Single Site Results.csv

**Supplemental Figure 6**. Model diagnostics QQ-plots for incident any impairment models.

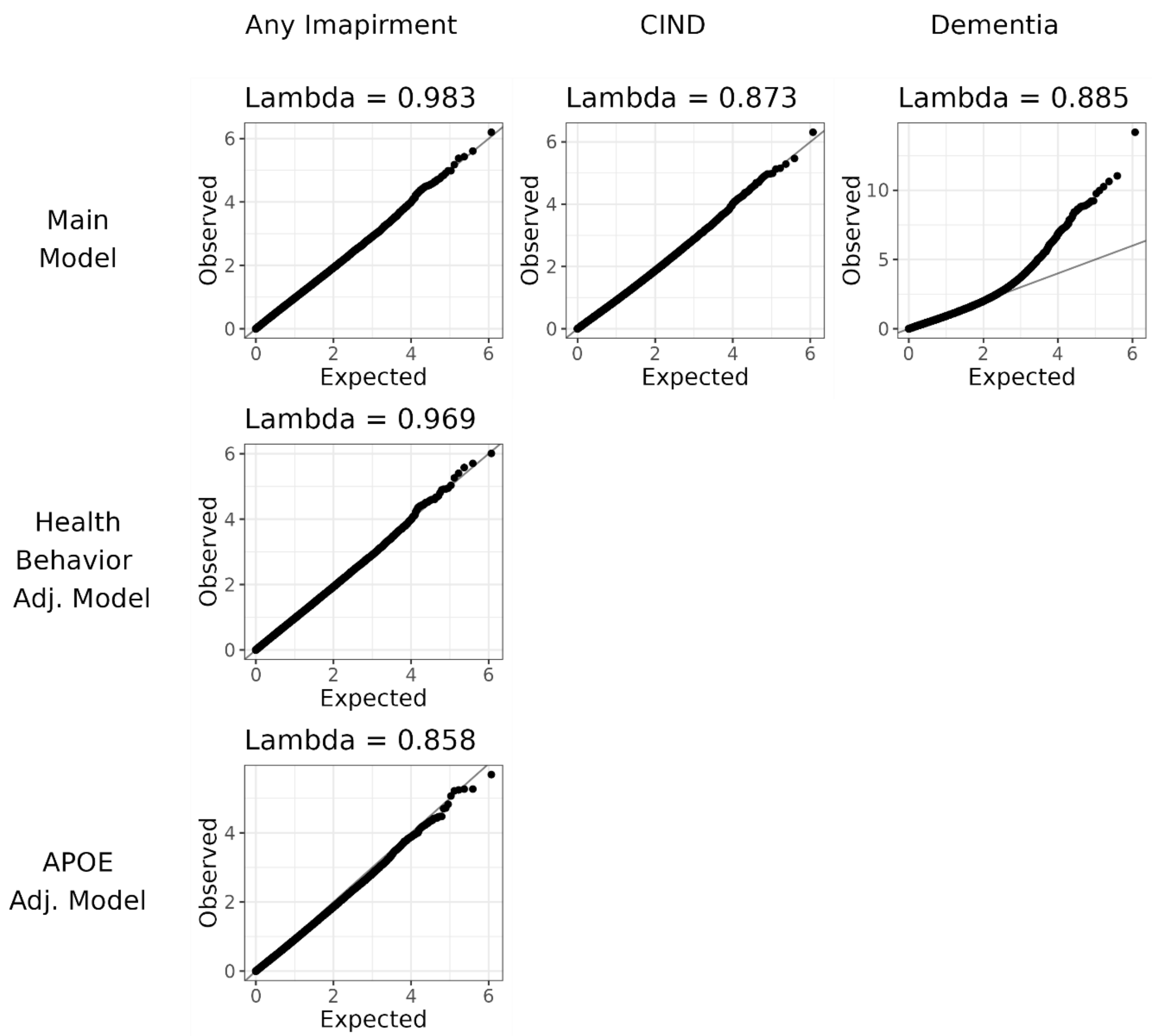

**Supplementary Table 14**. Gene ontology enrichment for prevalent CIND results.

File: Supplemental Table 14 Prevalent CIND Gene Ontology

| Pathway | Size | Enrichment Score | Normalized Enrichment Score | P-value | P-adj |
|---|---|---|---|---|---|
| ligand-gated monoatomic cation channel activity | 110 | 0.754317 | 1.17295 | 6.05E-06 | 0.00862 |
| ligand-gated channel activity | 147 | 0.733472 | 1.146254 | 1.06E-05 | 0.00862 |
| ligand-gated monoatomic ion channel activity | 146 | 0.7311 | 1.142503 | 1.88E-05 | 0.010191 |
| embryonic skeletal system development | 126 | 0.731382 | 1.140103 | 7.95E-05 | 0.032249 |
| skeletal system morphogenesis | 221 | 0.693315 | 1.090161 | 0.000239 | 0.07761 |
| spinal cord development | 102 | 0.737828 | 1.145756 | 0.000298 | 0.080689 |
| protein polyubiquitination | 217 | 0.693481 | 1.090054 | 0.000426 | 0.095634 |
| sex differentiation | 274 | 0.680295 | 1.072453 | 0.000472 | 0.095634 |
| reproductive structure development | 292 | 0.679777 | 1.072526 | 0.000563 | 0.101514 |
| reproductive system development | 296 | 0.679736 | 1.07268 | 0.000646 | 0.104733 |
| regulation of cell killing | 105 | 0.726778 | 1.129424 | 0.000746 | 0.110067 |
| renal system development | 320 | 0.673865 | 1.064183 | 0.000966 | 0.13063 |
| ubiquitin-like protein transferase activity | 439 | 0.665353 | 1.053848 | 0.001077 | 0.134362 |
| regulation of extent of cell growth | 108 | 0.715871 | 1.112942 | 0.001776 | 0.160072 |
| embryonic limb morphogenesis | 116 | 0.712781 | 1.109656 | 0.001684 | 0.160072 |
| embryonic appendage morphogenesis | 116 | 0.712781 | 1.109656 | 0.001684 | 0.160072 |
| regulation of axonogenesis | 155 | 0.697974 | 1.091823 | 0.001592 | 0.160072 |
| male gonad development | 144 | 0.69808 | 1.090907 | 0.001445 | 0.160072 |
| amine transport | 104 | 0.717538 | 1.114834 | 0.00209 | 0.16503 |
| polyol metabolic process | 101 | 0.715777 | 1.111023 | 0.002645 | 0.16503 |

**Supplementary Table 15**. Gene ontology enrichment for prevalent dementia results.

File: Supplemental Table 15 Prevalent Dementia Gene Ontology

| Pathway | Size | Enrichment Score | Normalized Enrichment Score | P-value | P-adj |
|---|---|---|---|---|---|
| protein serine kinase activity | 352 | 0.528036 | 1.278954 | 1.23E-09 | 2.00E-06 |
| protein serine/threonine kinase activity | 414 | 0.513165 | 1.247002 | 1.52E-08 | 1.23E-05 |
| Golgi vesicle transport | 275 | 0.531664 | 1.280676 | 5.93E-08 | 3.20E-05 |
| DNA-binding transcription activator activity | 462 | 0.49698 | 1.210453 | 1.28E-07 | 5.17E-05 |
| cell leading edge | 409 | 0.5021 | 1.219976 | 2.31E-07 | 6.24E-05 |
| DNA-binding transcription activator activity, RNA polymerase II-specific | 458 | 0.496908 | 1.210029 | 2.08E-07 | 6.24E-05 |
| regulation of anatomical structure size | 484 | 0.48841 | 1.19037 | 6.17E-07 | 0.000143 |
| cell-cell junction assembly | 155 | 0.555134 | 1.316924 | 2.87E-06 | 0.000377 |
| myeloid leukocyte activation | 232 | 0.521388 | 1.249648 | 2.91E-06 | 0.000377 |
| regulation of cellular component size | 349 | 0.504979 | 1.222814 | 3.04E-06 | 0.000377 |
| cell junction assembly | 440 | 0.490125 | 1.192813 | 2.46E-06 | 0.000377 |
| DNA-binding transcription factor binding | 461 | 0.486416 | 1.184695 | 2.80E-06 | 0.000377 |
| regulation of cytoskeleton organization | 480 | 0.485401 | 1.182571 | 3.25E-06 | 0.000377 |
| cell-cell junction | 493 | 0.482163 | 1.175439 | 2.91E-06 | 0.000377 |
| regulation of actin filament-based process | 351 | 0.497699 | 1.205467 | 3.77E-06 | 0.000407 |
| ubiquitin-like protein transferase activity | 439 | 0.483598 | 1.176787 | 4.11E-06 | 0.000417 |
| cellular response to abiotic stimulus | 321 | 0.501101 | 1.211348 | 6.19E-06 | 0.000558 |
| cellular response to environmental stimulus | 321 | 0.501101 | 1.211348 | 6.19E-06 | 0.000558 |
| postsynaptic density | 307 | 0.500992 | 1.209708 | 9.24E-06 | 0.000789 |
| carbohydrate homeostasis | 232 | 0.513789 | 1.231434 | 1.12E-05 | 0.000818 |

**Supplementary Table 16**. Gene ontology enrichment for incident any cognitive impairment CpGs.

File: Supplemental Table 16 Incident Any Impairment Gene Ontology.csv

| Pathway | Size | Enrichment Score | Normalized Enrichment Score | P-value | P-adj |
|---|---|---|---|---|---|
| pattern specification process | 454 | 0.688477 | 1.090747 | 7.28E-07 | 0.001013 |
| regionalization | 411 | 0.688361 | 1.089963 | 1.69E-06 | 0.001013 |
| embryonic organ development | 446 | 0.687073 | 1.088451 | 1.87E-06 | 0.001013 |
| cortical cytoskeleton | 102 | 0.765278 | 1.185806 | 4.25E-06 | 0.001603 |
| embryonic organ morphogenesis | 291 | 0.696884 | 1.099324 | 4.94E-06 | 0.001603 |
| cell cortex | 285 | 0.699502 | 1.103359 | 1.63E-05 | 0.004416 |
| regulation of neuron differentiation | 192 | 0.715288 | 1.121927 | 2.11E-05 | 0.004885 |
| muscle organ development | 333 | 0.687816 | 1.086766 | 2.44E-05 | 0.004954 |
| skeletal system morphogenesis | 221 | 0.708374 | 1.113508 | 2.78E-05 | 0.005008 |
| kidney development | 310 | 0.689845 | 1.089076 | 3.17E-05 | 0.005144 |
| anterior/posterior pattern specification | 213 | 0.707029 | 1.110529 | 4.18E-05 | 0.006166 |
| DNA-binding transcription activator activity | 462 | 0.673682 | 1.067474 | 7.05E-05 | 0.00953 |
| DNA-binding transcription activator activity, RNA polymerase II-specific | 458 | 0.672042 | 1.064781 | 0.00013 | 0.016266 |
| cytoplasmic ribonucleoprotein granule | 238 | 0.691333 | 1.087758 | 0.000212 | 0.02456 |
| ribonucleoprotein granule | 255 | 0.689007 | 1.085005 | 0.000239 | 0.02587 |
| actin filament binding | 202 | 0.697336 | 1.094427 | 0.000276 | 0.027949 |
| renal system development | 320 | 0.681864 | 1.076784 | 0.000303 | 0.028913 |
| morphogenesis of a branching structure | 200 | 0.693876 | 1.088799 | 0.000335 | 0.030181 |
| tRNA metabolic process | 167 | 0.699902 | 1.095097 | 0.000792 | 0.067632 |
| neuron migration | 170 | 0.70135 | 1.097734 | 0.00092 | 0.07465 |

**Supplemental Figure 7**. CpG island region enrichment of top (p-value < 0.01) CpGs for cross-sectional analyses.

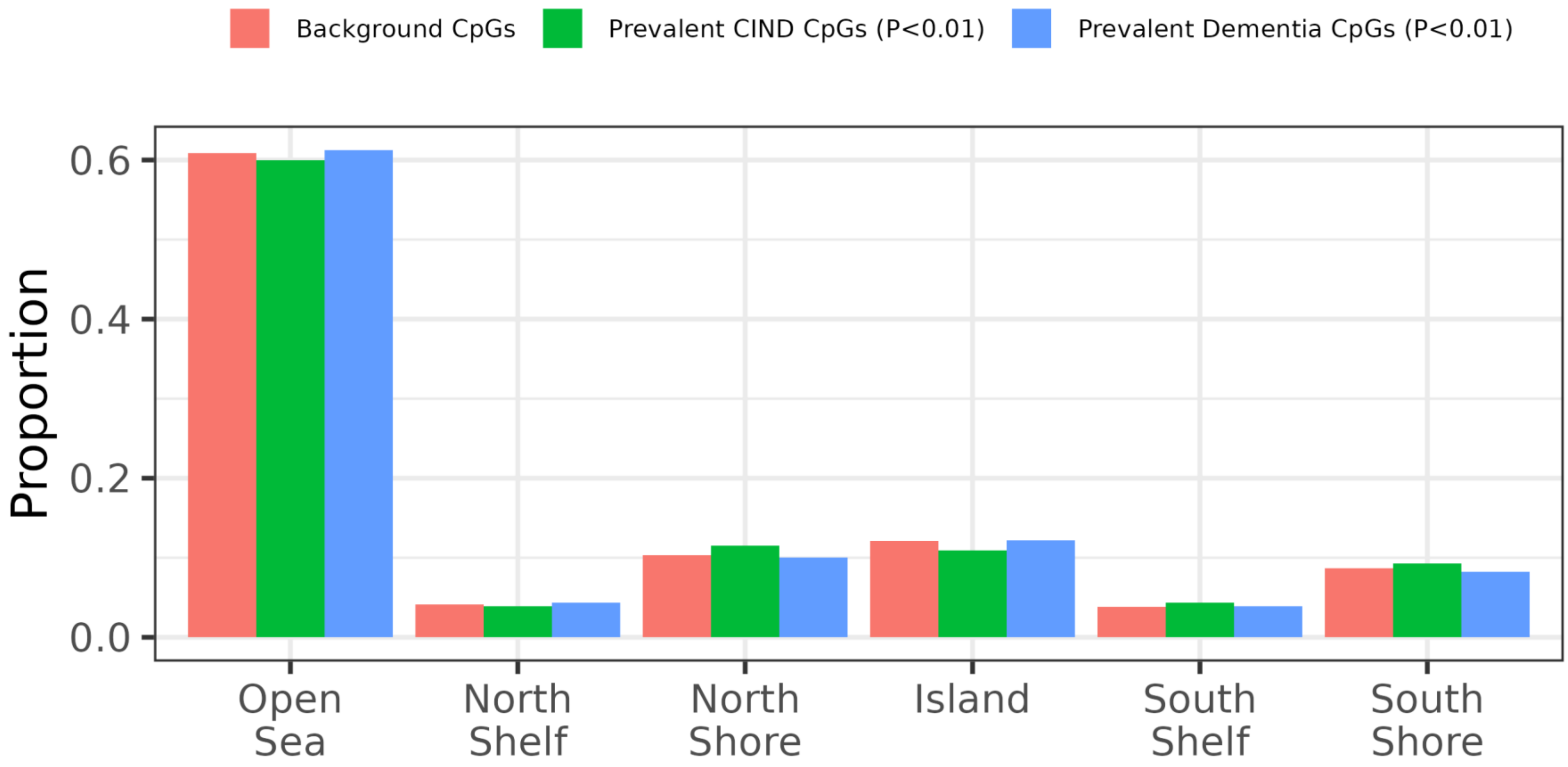

**Supplementary Figure 8**. CpG island region enrichment of top (p-value < 0.01) CpGs for incident analysis.

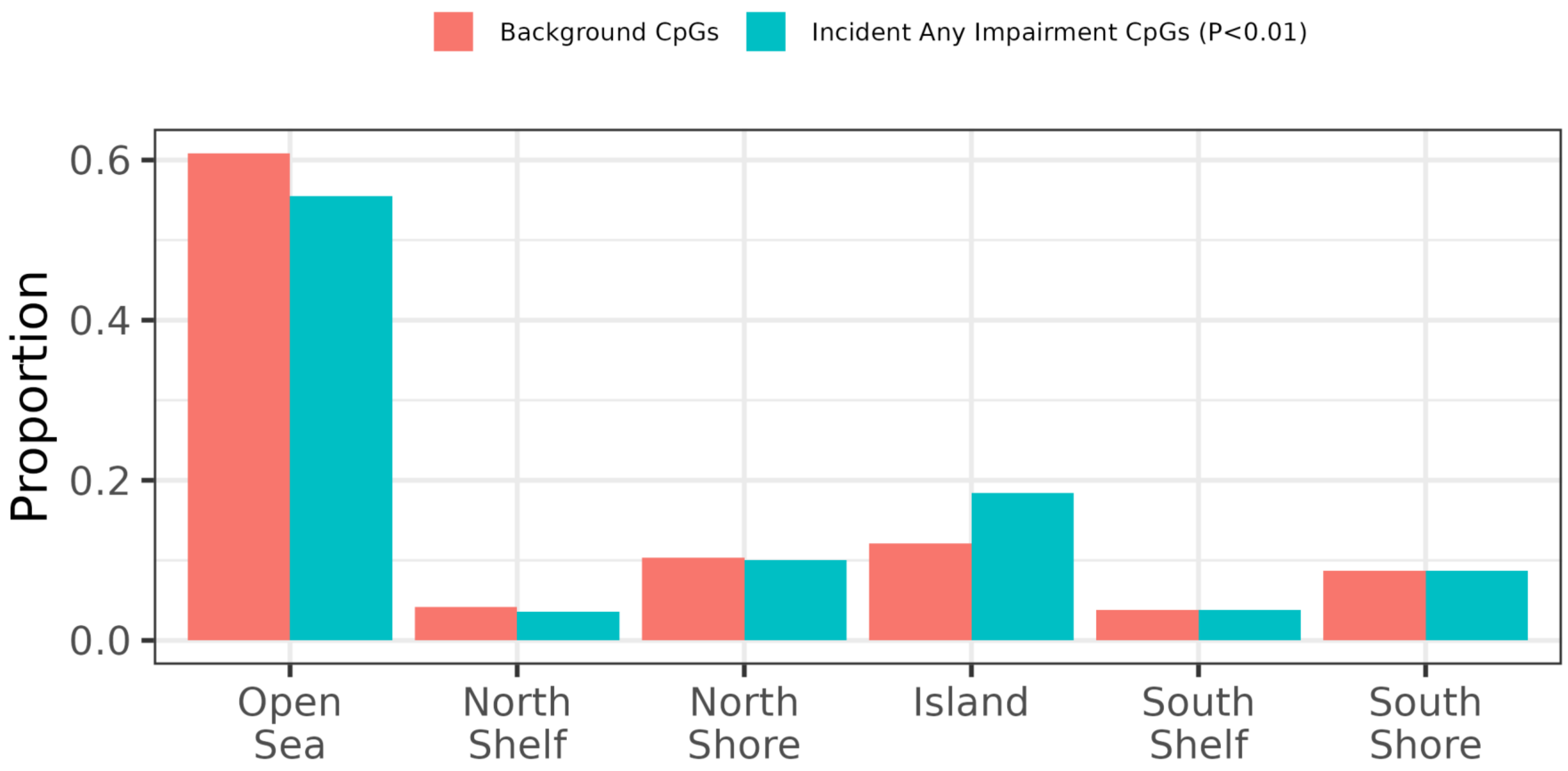

**Supplementary Figure 9**. eForge chromatin state enrichment for prevalent CIND top 1000 CpGs.

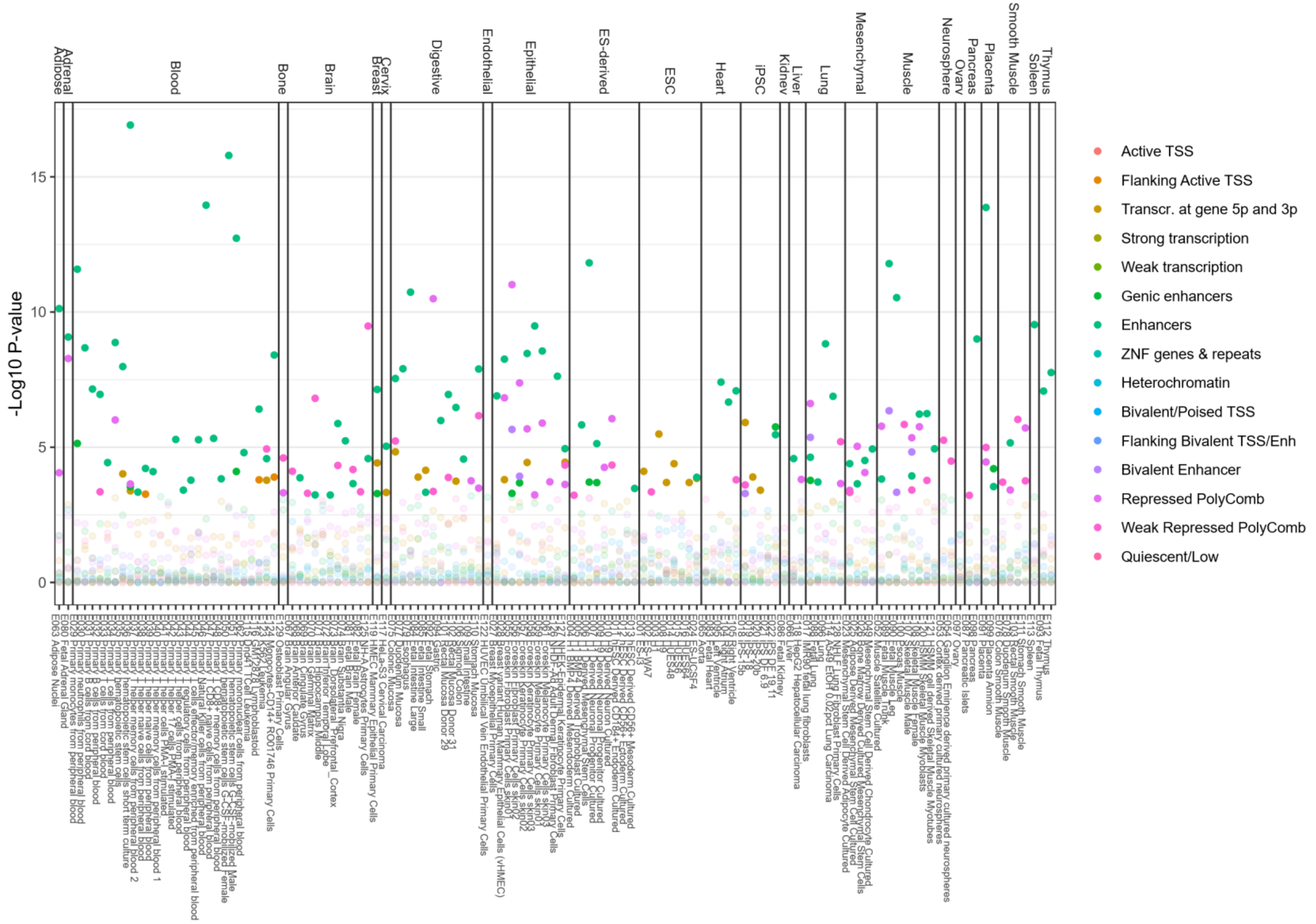

**Supplementary Figure 10**. eForge chromatin state enrichment for prevalent prevalent dementia top 1000 CpGs.

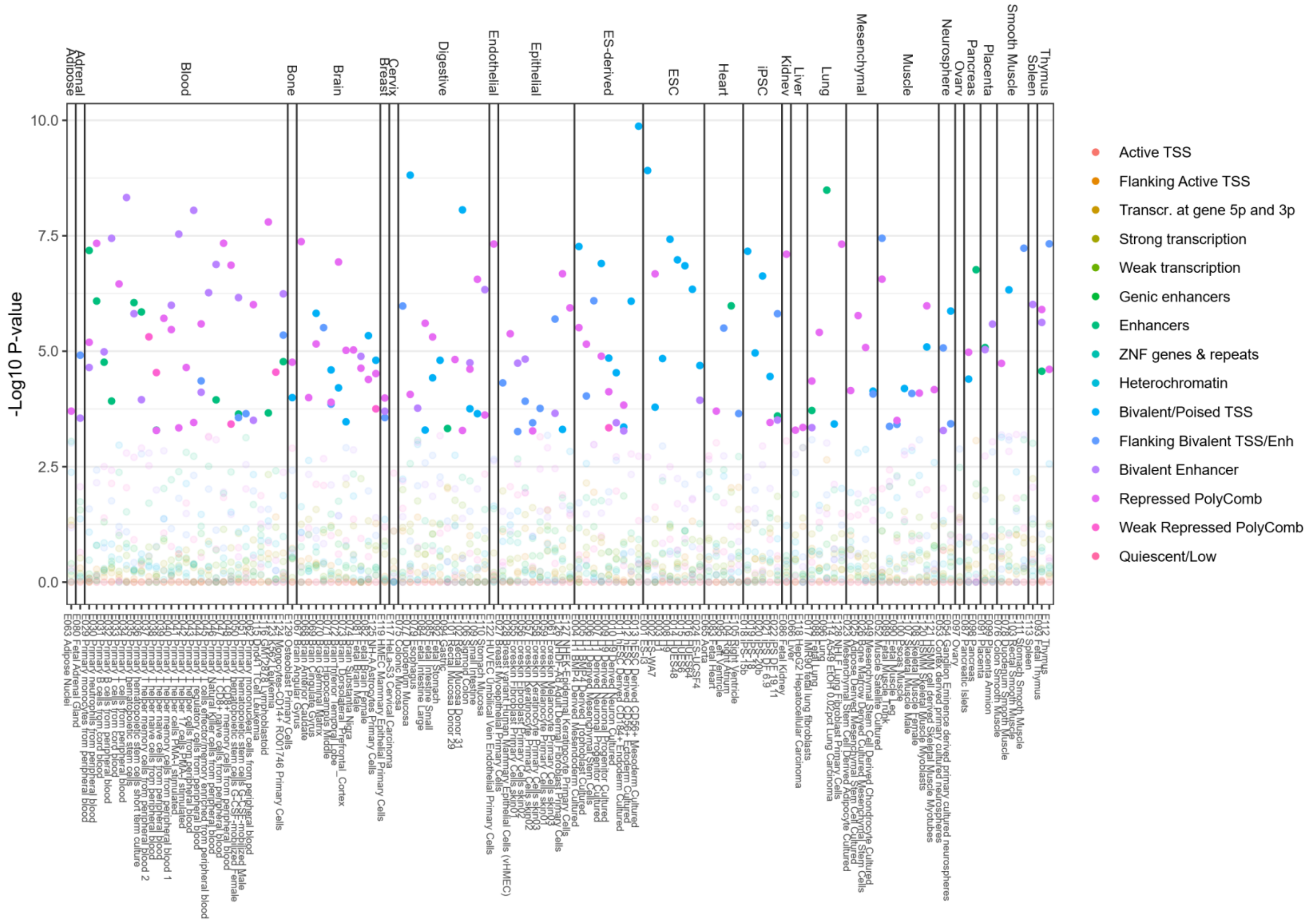

**Supplementary Figure 11**. eForge chromatin state enrichment for incident any cognitive impairment top 1000 CpGs.

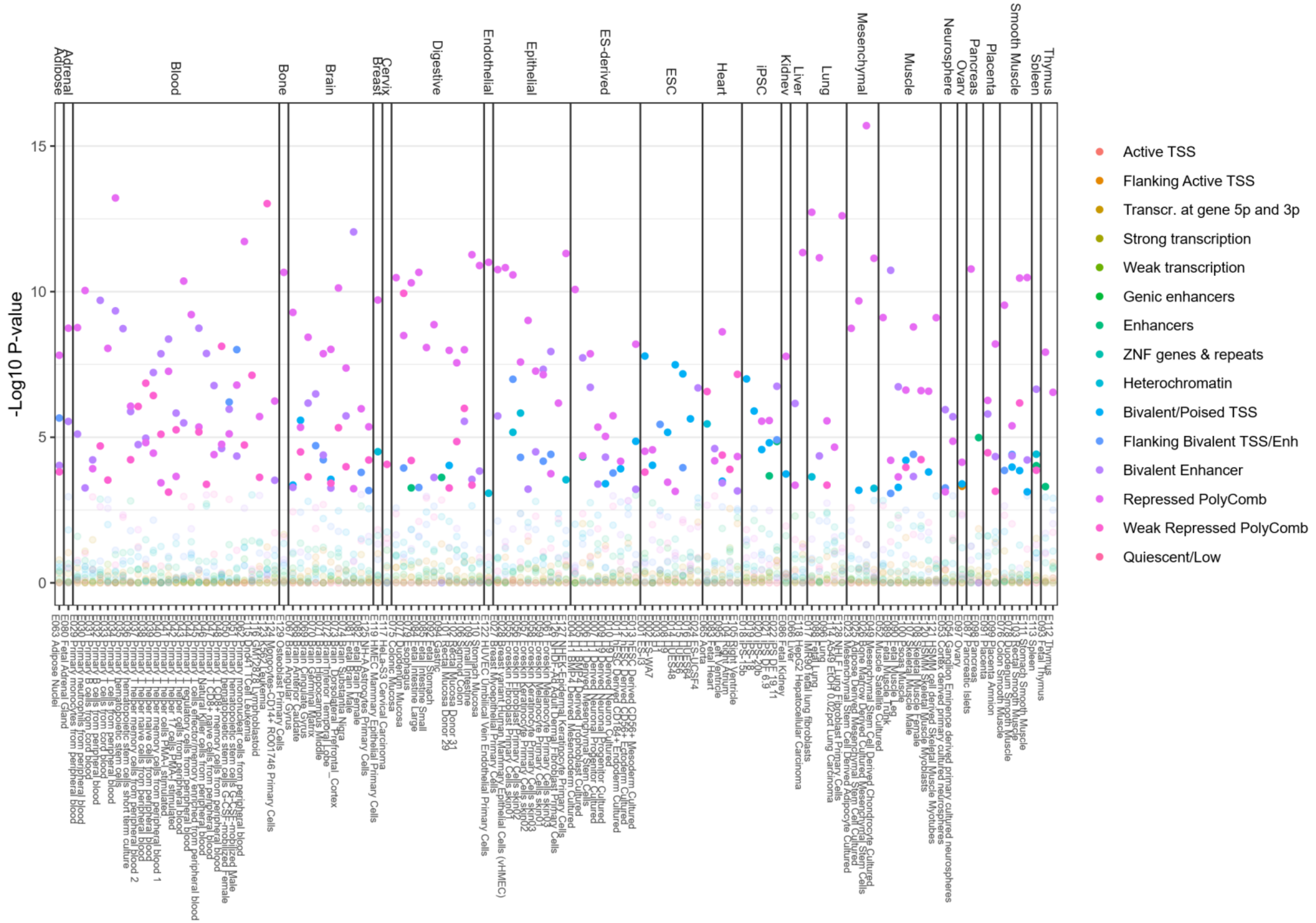

**Supplemental Figure 12.** Correlation of effect estimates for all prevalent analysis and incident analysis models.

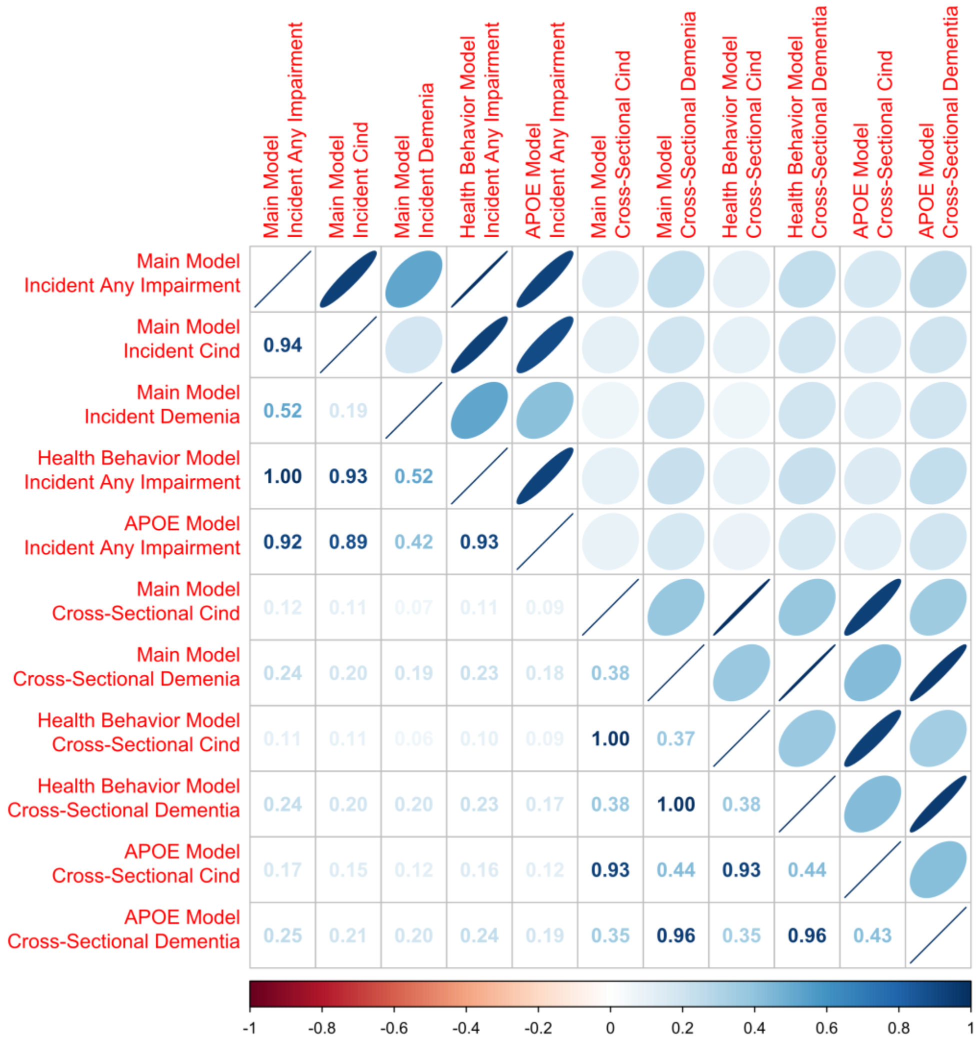

**Supplementary Figure 13**. Upset plot of overlap between HRS prevalent and incident cognitive impairment top CpGs (p-value < 0.01).

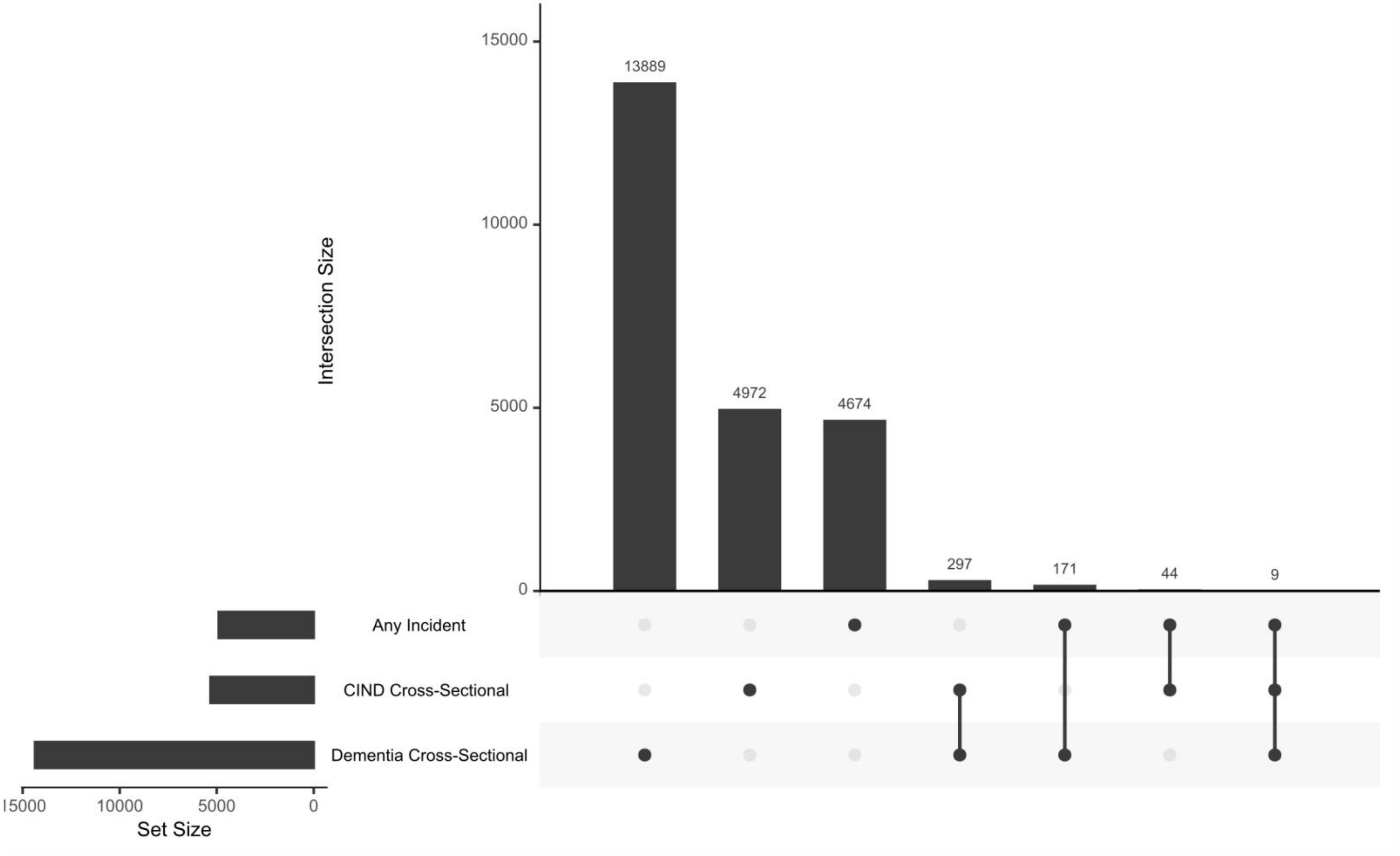

**Supplemental Figure 14.** Upset plot of overlap between HRS prevalent and incident cognitive impairment top CpGs (p-value < 0.01) with top 1000 CpGs related to mild cognitive impairment and top 1000 CpGs related to Alzheimer's disease in Roubroeks 2020 which examined 284 individuals (89 controls, 86 Alzheimer's disease, 109 mild cognitive impairment).

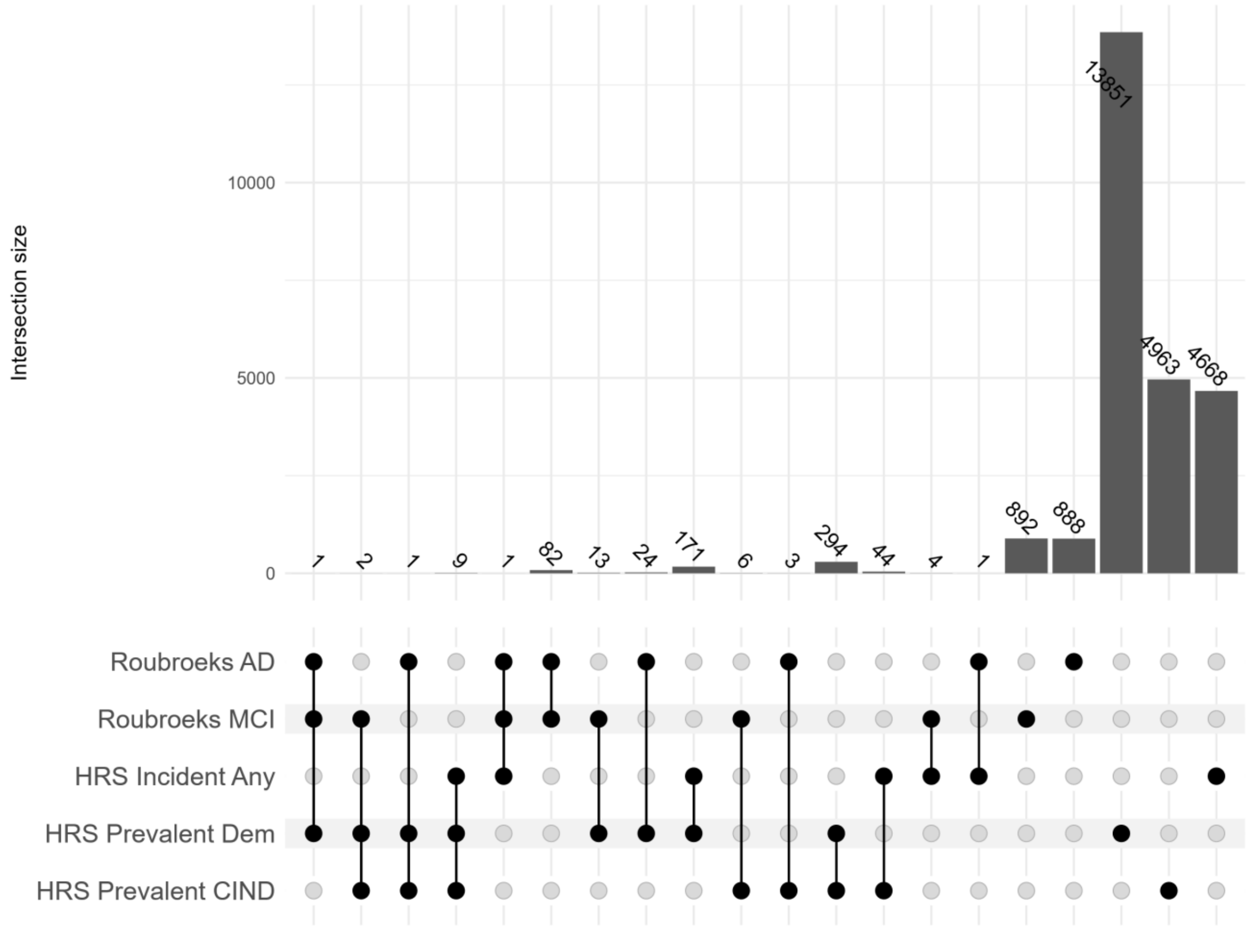

**Supplemental Figure 15.** Upset plot of overlap between HRS prevalent and incident cognitive impairment top CpGs (p-value < 0.01) with top CpGs (p-value < 0.01) in Sommerer 2022 which examined episodic memory in 1019 individuals cross-sectionally, and 626 individuals longitudinally.

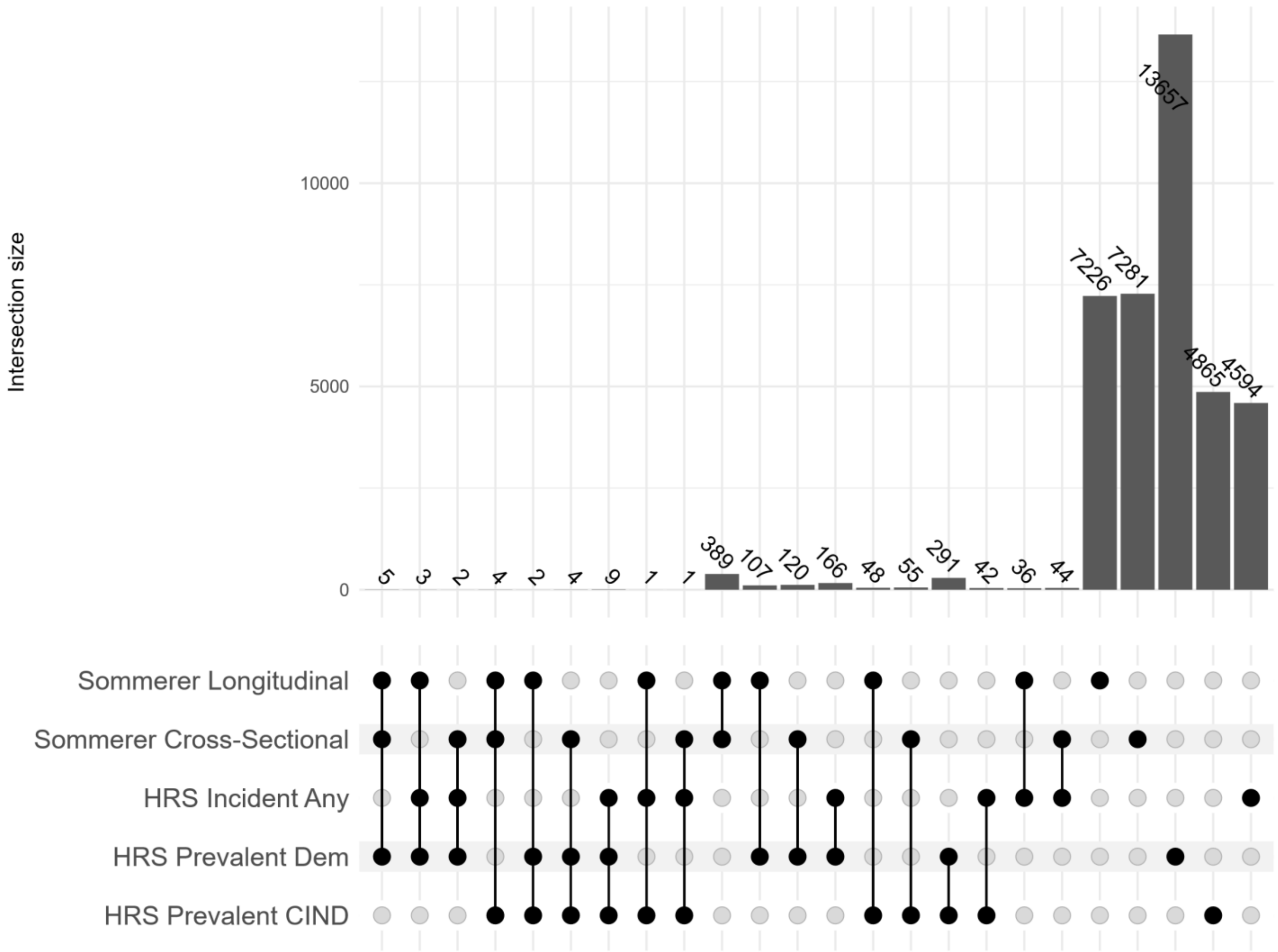
